# Association of the Liver and Plasma Lipidomes with the Histological Stage of Alcohol-Related Liver Disease

**DOI:** 10.1101/2021.07.13.21260429

**Authors:** Maja Thiele, Tommi Suvitaival, Kajetan Trošt, Min Kim, Andressa de Zewadzki, Maria Kjaergaard, Ditlev Nytoft Rasmussen, Katrine Prier Lindvig, Mads Israelsen, Sönke Detlefsen, Peter Andersen, Helene Bæk Juel, Trine Nielsen, Stella Georgiou, Vicky Filippa, Michael Kuhn, Suguru Nishijima, Lucas Moitinho-Silva, Peter Rossing, Jonel Trebicka, Ema Anastasiadou, Peer Bork, Torben Hansen, Cristina Legido Quigley, Aleksander Krag, on behalf of the MicrobLiver and GALAXY Consortia

**Author notes:** **Correspondence:**; Niels Steensens Vej 2-6, DK-2820 Gentofte, Denmark; and, Center for Liver Research, Department of Gastroenterology and Hepatology, Odense University Hospital, Odense, Denmark. These authors contributed equally to the work. Shared last authorship (Cristina Legido Quigley, Aleksander Krag). GALAXY consortium:* Ema Anastasiadou, Manimozhiyan Arumugam, Peer Bork, Torben Hansen, Jenny Presto, Hans Israelsen, Morten Karsdal, Cristina Legido-Quigley, Hans Olav Melberg, Maja Thiele, Jonel Trebicka, Aleksander Krag (coordinator). MicrobLiver consortium* Peer Bork, Mathias Mann, Jelle Matthijnssens, Aleksander Krag, Torben Hansen (coordinator). **Author contributions** AK, CLQ, MT and TH conceptualized and designed the study; MT, MKj, KPL, and MI recruited participants and performed the study investigations; SD scored the liver biopsies; DNR, KT and TS curated the data; KT and MKi conducted lipidomics analyses in liver and plasma; TS, KT, and CLQ did the formal analyses of lipidomics data; AK, CLQ, JT, KT, MKi, MT, TH and TS interpreted the results of the lipidomics analyses; SN, MKu, LMS and PB investigated the sphingolipid metabolism in the circulating transcriptome; EA, SG and VF investigated the sphingolipid metabolism in the circulating miRNA, TS carried out Mendelian randomization; MT, TS, KT, MKi, and CLQ drafted the manuscript; AK and TH acquired funds to perform the study. All authors revised the manuscript for important intellectual content and approved the final version.

## Abstract

**Background and aims:** Alcohol disturbs hepatic lipid synthesis and transport, but the role of lipid dysfunction in the severity of alcohol-related liver disease (ALD) is unclear. We therefore characterised the liver and plasma lipidome in a biopsy-controlled cohort of patients with early ALD.

**Methods:** We performed ultra-high-performance liquid chromatography quadrupole time-of-flight mass spectrometry for lipidomics of the liver and plasma from 315 patients, and of plasma from 51 healthy controls matched for age, gender and BMI. We correlated lipid levels with histological fibrosis, inflammation and steatosis, after correction for multiple testing and adjustment for age, gender, statin use, BMI, HbA1_c_, HOMA-IR, and ongoing drinking. Moreover, we investigated the mechanism of dysregulated sphingolipid metabolism by whole-blood transcriptomics and qPCR sequencing of miRNA.

**Results:** We detected 198 lipids in the liver and 236 lipids in the circulation from 18 lipid classes. Nearly all ceramides, sphingomyelins and lyso-phosphocholines in plasma decreased as fibrosis progressed. This was paralleled by a comparable decrease in the liver. Circulating and liver sphingomyelins were also inversely associated with hepatic inflammation. The lipidomic signature of healthy controls was only comparable to ALD patients with no fibrosis. Three circulating miRNA, highly involved in sphingomyelin metabolism, were dysregulated together with the mRNA expression of enzymes in the sphingomyelin degradation pathway. Mendelian randomization in Finnish and UK population biobanks externally validated our findings, suggesting a causal relationship between genetic disposition to ALD and low sphingolipid levels.

**Conclusions:** Liver fibrosis severity in alcohol-related liver disease is characterized by selective lipid depletion in blood and liver, indicating profound effects of progressive disease on the bioactive sphingolipids, already from early stages of fibrosis.

**Visual Abstract:** 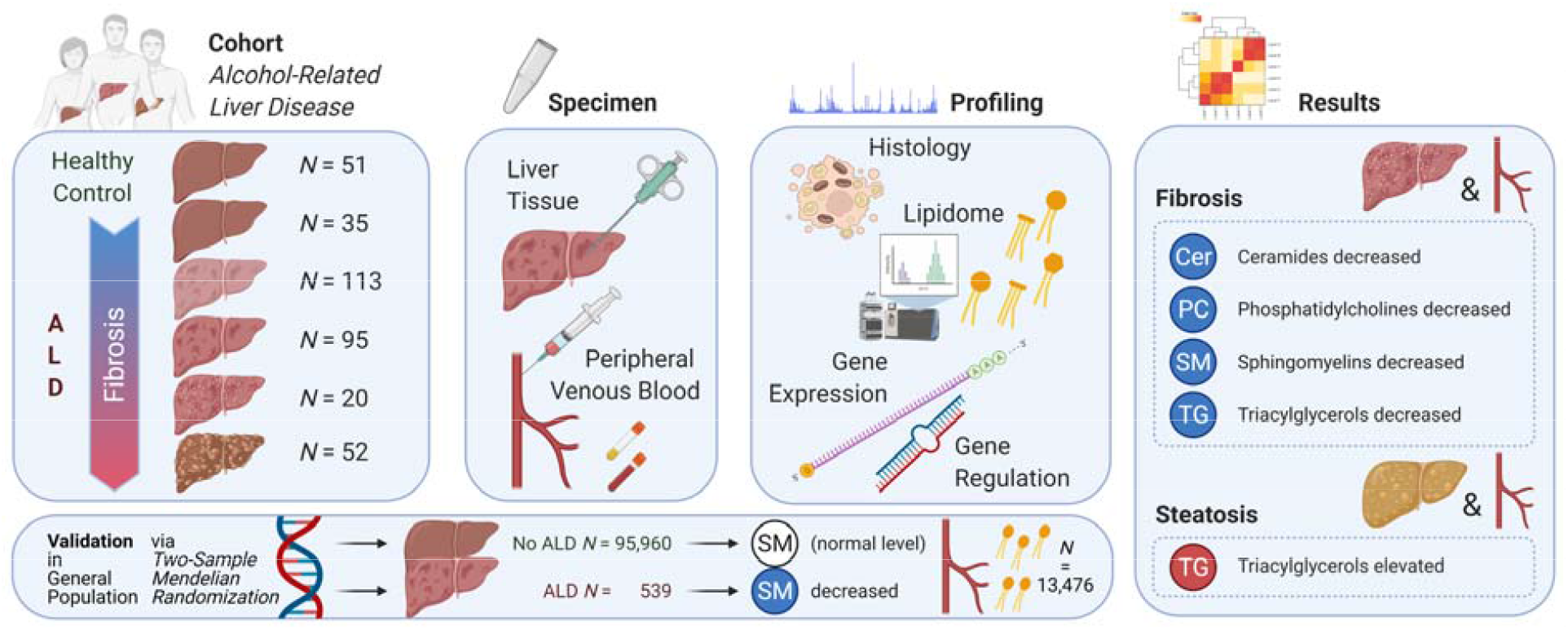

**Highlights:**

- The lipidome in the liver and circulation in alcohol-related liver disease (ALD)
- Sphingolipid, phospholipid and triacylglycerol levels were lowered in fibrosis
- Expression and regulation of genes in the sphingolipid pathway were affected
- ALD has a causal link to lowered sphingomyelin levels in blood

**Lay summary:** Alcohol causes a dysfunctional fat metabolism in the liver. In this study, we detected 198 different types of lipids in the liver and 236 in the blood stream of patients with different severity of alcohol-related liver disease. We found that patients with more severe scarring (fibrosis) of the liver, and more severe liver inflammation, had lower levels of sphingolipids both in the circulation and the liver. Sphingolipids regulate cell survival and inflammation, so they may be involved in the mechanism of progressive alcohol-related liver disease.

## INTRODUCTION

Alcohol is the leading cause of cirrhosis worldwide, with half of all global deaths from cirrhosis attributed to excess drinking [1]. One in five people with a harmful use of alcohol exhibit progressive liver fibrosis [2, 3]. The molecular basis of the development and progression of alcohol-related liver disease (ALD) is largely unknown [4, 5]. Hence, there is an urgent need to advance our understanding of the biochemical and pathophysiological changes in the liver and circulation during the progression of ALD.

Alcohol-induced lipid dysfunction is well known, in part driven by stress to the endoplasmic reticulum, which controls lipid and cholesterol synthesis [6, 7]. Consequently, alcohol impairs hepatocyte fatty acid oxidation and lipid transport, and increases lipogenesis, resulting in steatosis [7]. Furthermore, exposure to alcohol leads to hepatocellular stress, resulting in caspase activation, degradation of the cytoskeleton and subsequent apoptosis and ballooning, a hallmark of steatohepatitis [8, 9].

Triglycerides and fatty acids represent the major lipid classes accumulating in hepatocytes during the formation of steatosis, but they are inert and not lipotoxic [10]. Considering that alcohol disrupts the global hepatic lipid metabolism, a comprehensive lipidomics catalogue is more appropriate [11]. Lipidomics studies of non-alcoholic fatty liver disease (NAFLD) indicate that bioactive lipids such as phosphocholines and sphingolipids, which control cell integrity and function, play a major role in lipotoxicity [12]. Human case-control studies in NAFLD have found that disease severity correlates with phosphocholines and sphingolipids, but results from these studies have been somewhat contradictory and none combined liver and plasma lipidomics [13-15]. Evidence is even more scarce in ALD, where it is limited to case-control studies in patients with alcohol-related cirrhosis and alcoholic hepatitis [16, 17].

There is consequently a need for investigations into the global lipidome in, both, circulation and liver tissue across the spectrum of alcohol-related liver fibrosis. We aimed to describe differences in the levels of individual lipids across lipid classes in plasma and liver tissue according to liver fibrosis stage, inflammation grade and steatosis score in a cohort study of biopsy-controlled patients with alcohol-related liver disease, compared to matched healthy controls. Finally, we validated our key findings by investigating the expression and regulation of genes involved in sphingolipid metabolism, together with conducting a Mendelian randomization study.

## METHODS

We conducted a cross-sectional cohort study of patients with ALD, and matched, healthy control subjects. Both studies received approval by the Danish data protection agency (13/8204, 16/3492) and the ethics committee for Region of Southern Denmark (ethical ID S-20120071, S-20160021, S-20170087, S-20160006G). The ALD cohort has been described in detail in two previously published studies [18, 19]. All participants gave written, informed consent prior to inclusion. Here, we describe the research methods in overview. Further details can be found in the Supplementary Material.

### Patients

We recruited consecutive patients with a history of chronic, excessive, alcohol use for at least one year (>14 units/week for women and >21 units/week for men). Additional inclusion criteria were age 18-75 years and informed consent to undergo a liver biopsy. We excluded patients in case of decompensated cirrhosis assessed by ultrasonography or endoscopy; competing liver disease; alcoholic hepatitis; mechanical bile duct obstruction; any debilitating disease with an expected survival of less than one year; disseminated malignancy; or inability to comply with the study protocol.

### Healthy controls

We matched 51 healthy controls with the ALD patients based on gender, age and body mass index (BMI). Exclusion criteria for healthy controls were prior excessive use of alcohol, any daily drinking, average weekly alcohol intake exceeding seven units for women and 14 units for men, binge drinking (≥5 units at a time), known liver disease of any etiology, use of illegal substances, any medication other than occasional mild pain relievers, or any chronic disease either self-reported, discovered during inclusion, or occurring within six months after inclusion.

### Study investigations

All investigations were performed on the same day, after a 10-minute rest, preceded by an overnight fast. Investigations included standardized questionnaires regarding medical history, current and prior alcohol history, and medications. We measured blood pressure, BMI, glycosylated hemoglobin A1c (HbA1c), plasma triglycerides, cholesterol, high- and low-density lipoprotein, and calculated the homeostatic model assessment of insulin resistance (HOMA-IR). We categorised patients as abstinent if they did not consume alcoholic beverages for at least seven days leading up to inclusion.

Finally, we performed percutaneous liver biopsy in ALD patients. One pathologist evaluated biopsy quality (≥10mm length and ≥5 portal tracts or presence of regenerative nodules), scored fibrosis stage according to the Pathology Committee of the NASH Clinical Research Network (NAS-CRN; Kleiner F0-F4), and used the NAS-CRN Activity Score for semi-quantitative evaluation of macro-vesicular steatosis (S0-S3), hepatocyte ballooning (B0-B2) and lobular inflammation (I0-I3) [20]. For a composite measure of inflammatory activity, we used the sum of lobular inflammation and ballooning (0-5).

### Plasma lipidomics

We performed untargeted lipidomics on 500μL lithium-heparin plasma, centrifuged at 2000G and stored at -80°C immediately after sampling. We used internal standards according to protocols at Steno Diabetes Center Copenhagen [21]. We identified lipids by ultra-high-performance liquid chromatography (1290 Infinity UHPLC system) coupled to quadrupole time-of-flight mass spectrometry (Agilent 6550 Q-ToF) from Agilent Technologies (Santa Clara, California, USA), which separates molecules based on their polarity, after which lipids can be classified based on their size and degree of unsaturation [22]. The untargeted lipidomics method has previously been validated across 31 European laboratories using the National Institute of Standards and Technology Standard Reference Material of human plasma metabolites [23].

### Liver lipidomics

We prepared liver biopsy samples according to previously established methods, modified to suit the experimental design and analytical instruments [24]. We used 5 mm of liver tissue, snap-frozen in liquid nitrogen immediately after biopsy. Each tissue sample were transferred from -80°C into Covaris TT05 tissue tubes, immerged into liquid nitrogen, and crushed with Covaris CryoPrep impactor CP02. Finally, two mg of the powdered liver tissue underwent chloroform-methanol extraction, with the exact mass used for normalization. Finally, we performed identical instrumental analysis as for plasma lipidomics. We performed quality control of individual measurements using lipidomics of pooled extracts as standard, and tested liver sample preparation variability using normal mouse liver tissue.

### Pre-processing, quality control and post-processing of lipidomics data

We used MZmine 2 to preprocess the lipidomics data, and a pipeline in R for quality control and post-processing. To denoise the data from technical variation, we normalized the peak areas of lipid measurements by the peak areas of internal standards, and batch corrected the values. Lipids with over 20 % missing values were omitted from subsequent analysis, and remaining missing values imputed using the k-nearest neighbor algorithm. We standardized all data variables to zero mean and unit variance for the inference of standardized effect sizes.

### Sphingolipid regulation

Post-hoc, we investigated the expression and regulation of the sphingomyelin degradation and synthesis pathways from whole-blood mRNA and miRNA analyses. Moreover, we tested the causality and its specificity between ALD and sphingomyelin levels via Mendelian randomization meta-analysis in the FinnGen and UK biobanks. These analyses are described in Supplementary Methods.

### Data analysis

First, we tested liver and plasma lipids separately, correlating each lipid species to fibrosis stage, using analysis of covariance (ANCOVA) coupled to pairwise post-hoc tests. To control for potential confounders, we adjusted all ANCOVA regression models for gender, age, BMI, HbA1_c_, HOMA-IR, systolic blood pressure, use of statins, and abstinence from alcohol at the time of inclusion.

We selected those lipid species where the ANCOVA showed a significant association with fibrosis to proceed to pairwise analyses of fibrosis stages, using no liver fibrosis (F0) as the reference. Moreover, we performed identical analyses for the inflammation grade and steatosis score. Six heatmaps graphically represent those analyses: three for the plasma lipidome and its associations with increasing stage of fibrosis, inflammation, and steatosis; and three for the liver lipidome. We used chord diagrams [25] to integrate all these associations and conducted enrichment analyses of lipid classes.

Finally, we investigated lipids that simultaneously were associated with disease severity both in the liver and in plasma: We identified the doubly-associated lipid species with the strongest early associations with fibrosis, inflammation and steatosis, and visualized the levels of these three lipid species with violin plots.

We corrected all results for multiple testing over the lipidome using the Benjamini-Hochberg method. We analysed the data with R (version 3.6.2) using the lipidomeR [26], circlize, gplots, and ggplot2 packages. For more details, see Supplementary Material.

## RESULTS

### Patients and healthy controls

We included 315 patients with ALD and 51 matched healthy controls between 2013 and 2018 (Table 1). We assessed steatosis, ballooning, and lobular inflammation in liver tissue from 312 ALD participants, whereas three biopsies showed regenerative nodules diagnostic of cirrhosis, but not enough tissue for reliable inflammation or steatosis scoring. We acquired a lipidomic profile of the liver in 301 of the participants, whereas 10 biopsies were used for adjustment of the method, and four liver samples did not contain enough material for lipidomics.

**Table 1:**
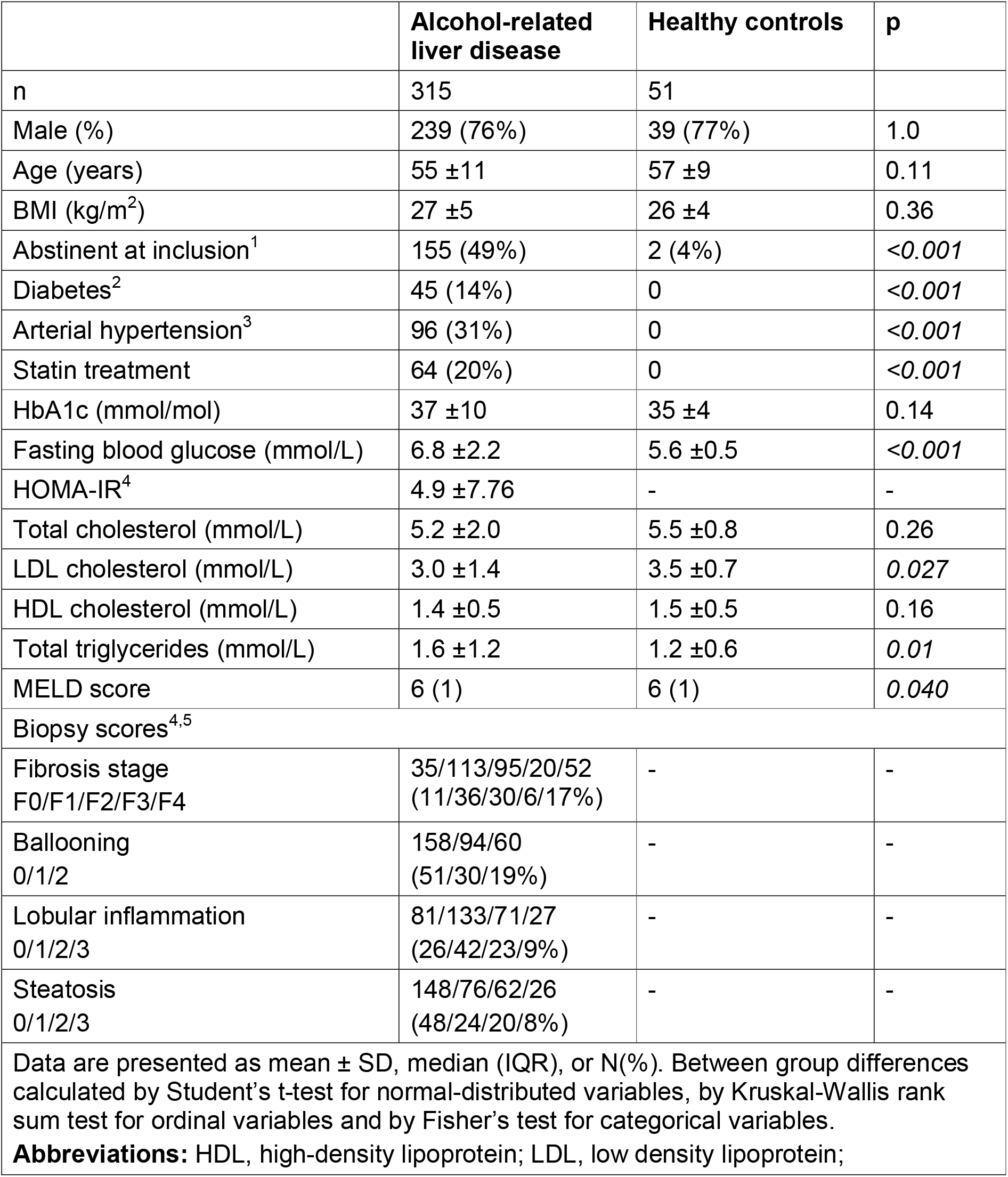
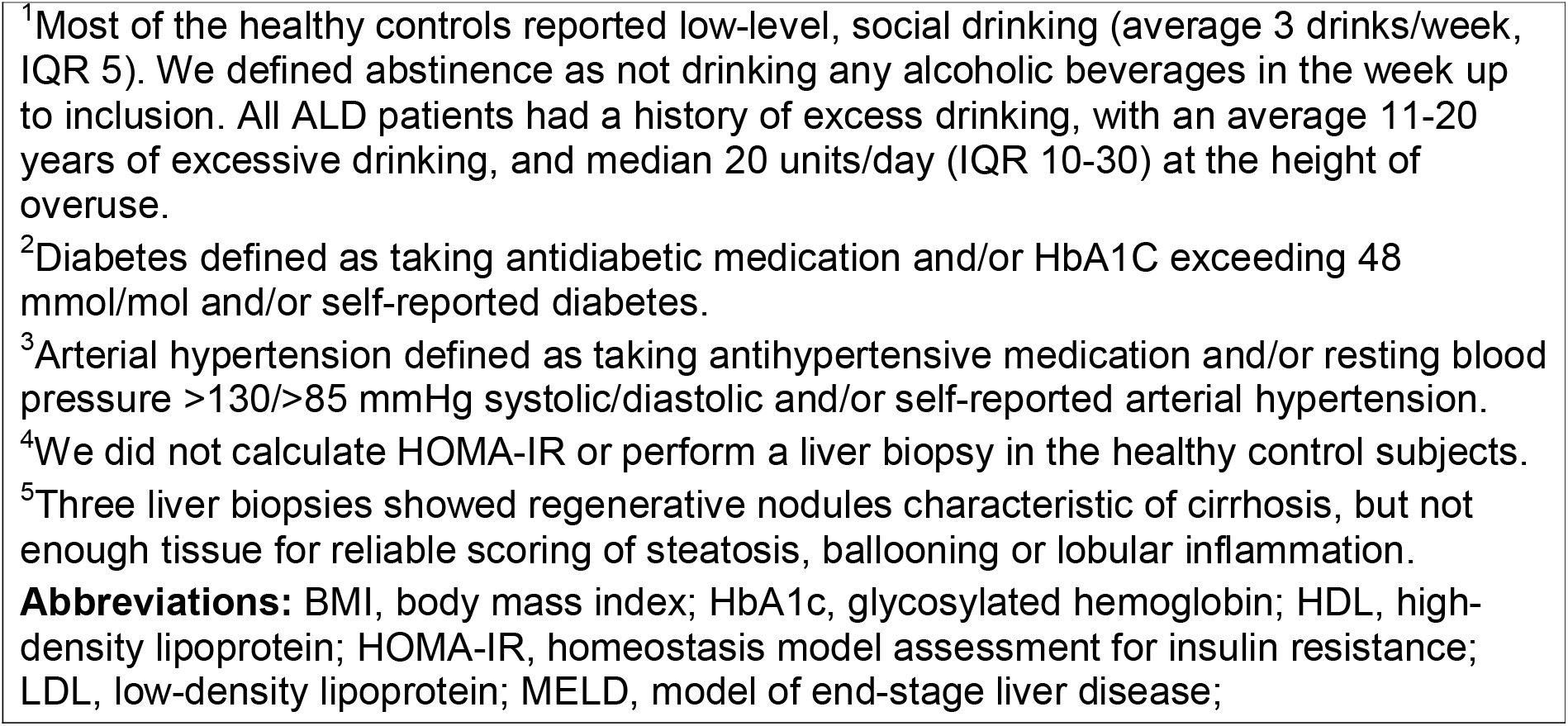
Characteristics of 315 patients with alcohol-related liver disease (ALD) and 51 healthy controls matched for age, gender and BMI.

|  | Alcohol-related liver disease | Healthy controls | p |
| --- | --- | --- | --- |
| n | 315 | 51 |  |
| Male (%) | 239 (76%) | 39 (77%) | 1.0 |
| Age (years) | 55 ±11 | 57 ±9 | 0.11 |
| BMI (kg/m<sup>2</sup>) | 27 ±5 | 26 ±4 | 0.36 |
| Abstinent at inclusion<sup>1</sup> | 155 (49%) | 2 (4%) | <0.001 |
| Diabetes<sup>2</sup> | 45 (14%) | 0 | <0.001 |
| Arterial hypertension<sup>3</sup> | 96 (31%) | 0 | <0.001 |
| Statin treatment | 64 (20%) | 0 | <0.001 |
| HbA1c (mmol/mol) | 37 ±10 | 35 ±4 | 0.14 |
| Fasting blood glucose (mmol/L) | 6.8 ±2.2 | 5.6 ±0.5 | <0.001 |
| HOMA-IR<sup>4</sup> | 4.9 ±7.76 | - | - |
| Total cholesterol (mmol/L) | 5.2 ±2.0 | 5.5 ±0.8 | 0.26 |
| LDL cholesterol (mmol/L) | 3.0 ±1.4 | 3.5 ±0.7 | 0.027 |
| HDL cholesterol (mmol/L) | 1.4 ±0.5 | 1.5 ±0.5 | 0.16 |
| Total triglycerides (mmol/L) | 1.6 ±1.2 | 1.2 ±0.6 | 0.01 |
| MELD score | 6 (1) | 6 (1) | 0.040 |
| Biopsy scores<sup>4,5</sup> |  |  |  |
| Fibrosis stage<br>F0/F1/F2/F3/F4 | 35/113/95/20/52<br>(11/36/30/6/17%) | - | - |
| Ballooning<br>0/1/2 | 158/94/60<br>(51/30/19%) | - | - |
| Lobular inflammation<br>0/1/2/3 | 81/133/71/27<br>(26/42/23/9%) | - | - |
| Steatosis<br>0/1/2/3 | 148/76/62/26<br>(48/24/20/8%) | - | - |
| Data are presented as mean ± SD, median (IQR), or N(%). Between group differences calculated by Student's t-test for normal-distributed variables, by Kruskal-Wallis rank sum test for ordinal variables and by Fisher's test for categorical variables.<br>**Abbreviations:** HDL, high-density lipoprotein; LDL, low density lipoprotein; |  |  |  |

<sup>1</sup>Most of the healthy controls reported low-level, social drinking (average 3 drinks/week, IQR 5). We defined abstinence as not drinking any alcoholic beverages in the week up to inclusion. All ALD patients had a history of excess drinking, with an average 11-20 years of excessive drinking, and median 20 units/day (IQR 10-30) at the height of overuse.
<sup>2</sup>Diabetes defined as taking antidiabetic medication and/or HbA1C exceeding 48 mmol/mol and/or self-reported diabetes.
<sup>3</sup>Arterial hypertension defined as taking antihypertensive medication and/or resting blood pressure >130/>85 mmHg systolic/diastolic and/or self-reported arterial hypertension.
<sup>4</sup>We did not calculate HOMA-IR or perform a liver biopsy in the healthy control subjects.
<sup>5</sup>Three liver biopsies showed regenerative nodules characteristic of cirrhosis, but not enough tissue for reliable scoring of steatosis, ballooning or lobular inflammation.
**Abbreviations:** BMI, body mass index; HbA1c, glycosylated hemoglobin; HDL, high-density lipoprotein; HOMA-IR, homeostasis model assessment for insulin resistance; LDL, low-density lipoprotein; MELD, model of end-stage liver disease;

We detected 198 lipids in the liver and 236 lipids in the circulation, coming from 18 lipid families or classes: ceramides (Cer), diacylglycerols (DG), free fatty acids (FA) hexocyl-ceramides (HexCer), lactocyl-ceramides (LacCer); lyso-phosphocholines (LPC), alkyl or alkenyl ether LPCs (LPC-O/Ps), lyso-phosphatidylethanolamines (LPE), phosphocholines (PC), alkyl or alkenyl ether PC’s (PC-O/P), phosphatidylethanolamines (PE), alkyl or alkenyl ether PEs (PE-O/P), phosphatidylglycerols (PG), phosphatidylinositols (PI), phosphatidylserines (PS), sulfatide HexCer (SHexCer), sphingomyelins (SM), and triacylglycerols (TG) (Supplementary Tables 1 and 2 for lists of all identified lipids).

### The plasma lipidome of alcohol-related liver disease patients compared to healthy, matched controls

The plasma lipidome of the healthy controls was similar to patients with no fibrosis (F0) with the exception of only two lipids (one FA and one PC, P<0.05 after Benjamini-Hochberg correction; Supplementary Figure S1). Further, 22 ALD patients did not have fibrosis, inflammation or steatosis on liver biopsy. These liver-healthy ALD patients had aberrated plasma levels of five phospholipids (P<0.05), when compared to healthy controls.

### Liver lipidome and fibrosis

We found aberrations in 13 lipid classes in the liver, associated with moderate fibrosis (F2), severe fibrosis (F3) and cirrhosis (F4), independent of gender, age, BMI, glucose metabolism, blood pressure, use of lipid-lowering medication, and abstinence from alcohol at the time of inclusion (Figure 1).

**Figure 1:**
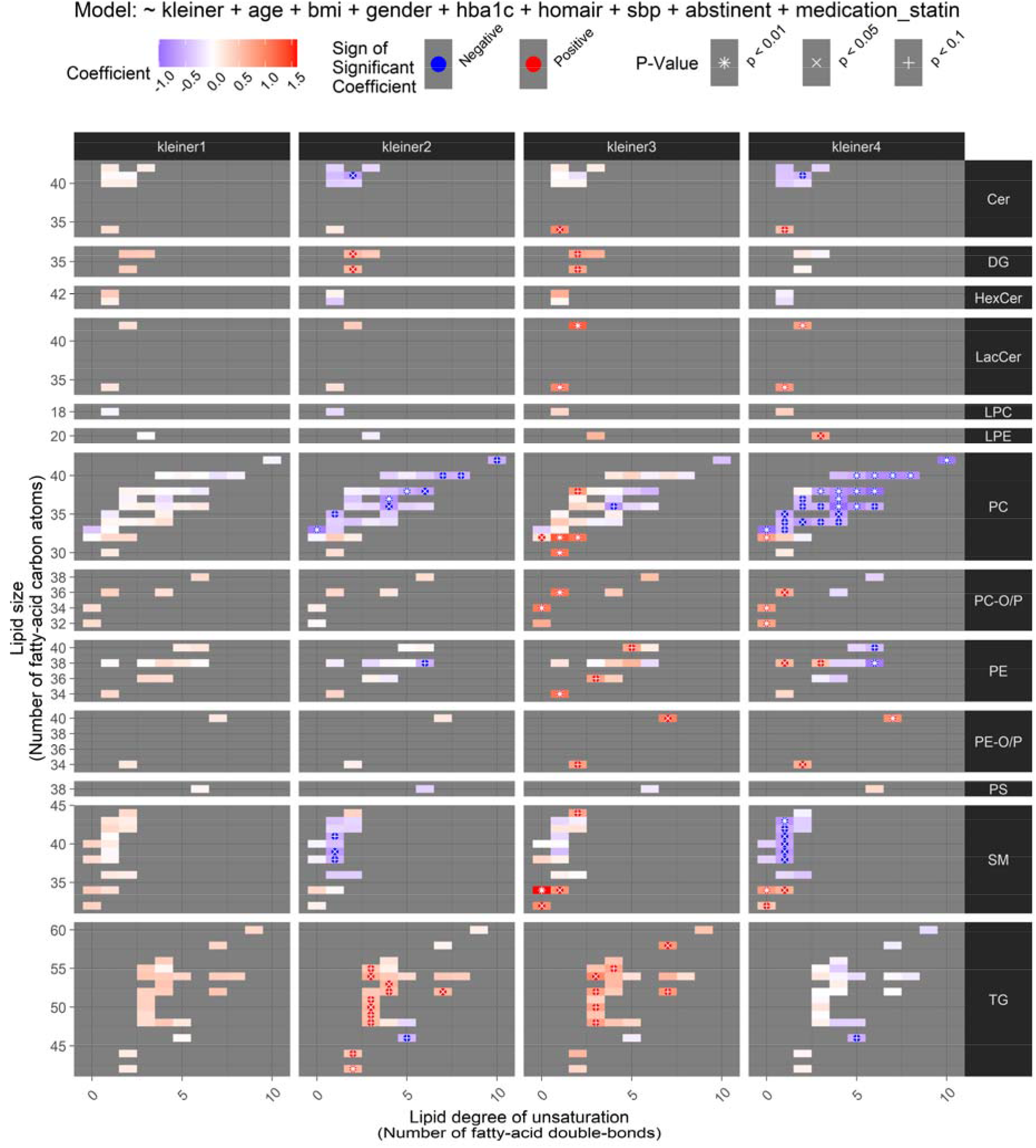
The liver lipidome according to fibrosis stage. Heatmaps of the liver lipidomic aberrations in alcohol-related liver fibrosis (stages as columns, Kleiner F0 as reference) are shown, after adjustment for possible confounders (age, sex, BMI, HbA1C, HOMA-IR, systolic blood pressure, abstinence from alcohol at the time of inclusion, and statin use). Liver lipids from 13 lipid classes (rows) are affected in fibrosis. For each lipid class, individual lipid species are organized by size (y-axis) and degree of unsaturation (x-axis). Each colored rectangle in a panel corresponds to one measured lipid species and the color of the rectangle shows the difference in the level of the lipid species between the fibrosis stage in question and a healthy liver (red is elevated, blue is reduced). Associations by ANCOVA with statistical significance after Benjamini-Hochberg correction for multiple testing are highlighted with asterisk (*), cross (x) and plus (+) signs, respectively, for p < 0.01, 0.05 and 0.1. **Abbreviations:** Cer, ceramides; DG, diacylglycerols; HexCer, hexocyl-ceramides; LacCer, lactocyl-ceramides; LPC, lyso-phosphocholines; LPE, lyso-phosphatidylethanolamines; PC, phosphocholines; PC-O/P, alkyl-acyl phosphocholines; PE, phosphatidylethanolamines; PE-O/P, alkyl or alkenyl ether phosphatidylethanolamines; PS, phosphatidylserines; SM, sphingomyelins; TG, triacylglycerols.

Notably, lipids belonging to PC and SM classes displayed an increasingly worsening depletion of the longer and unsaturated lipids, contrasted by an increase in the shorter and saturated lipids. We observed the most dysbalanced liver lipidome in cirrhosis patients, where a negative association for large and mid-sized PCs and SMs was contrasted by positive associations in small-sized, saturated or monounsaturated lipids from the classes Cer, LacCer, PC, PC-O/P, PE, and SM.

Five TGs and two DGs showed a positive correlation in patients with fibrosis stages F2-F3 but not in cirrhosis, indicating a loss of the excess acylglycerols when progressing from moderate and severe fibrosis to cirrhosis.

### Plasma lipidome and fibrosis

In plasma, we found qualitatively similar but stronger associations as in the liver (Figure 2). We observed strong, negative associations for several Cers and SMs. Nearly all lipids from both these sphingolipid classes showed signs of decreasing levels already at minimal fibrosis (Supplementary Figure S1). This negative association strengthened with increasing stage of fibrosis, rendering the entire SM and Cer classes significant at cirrhosis.

**Figure 2:**
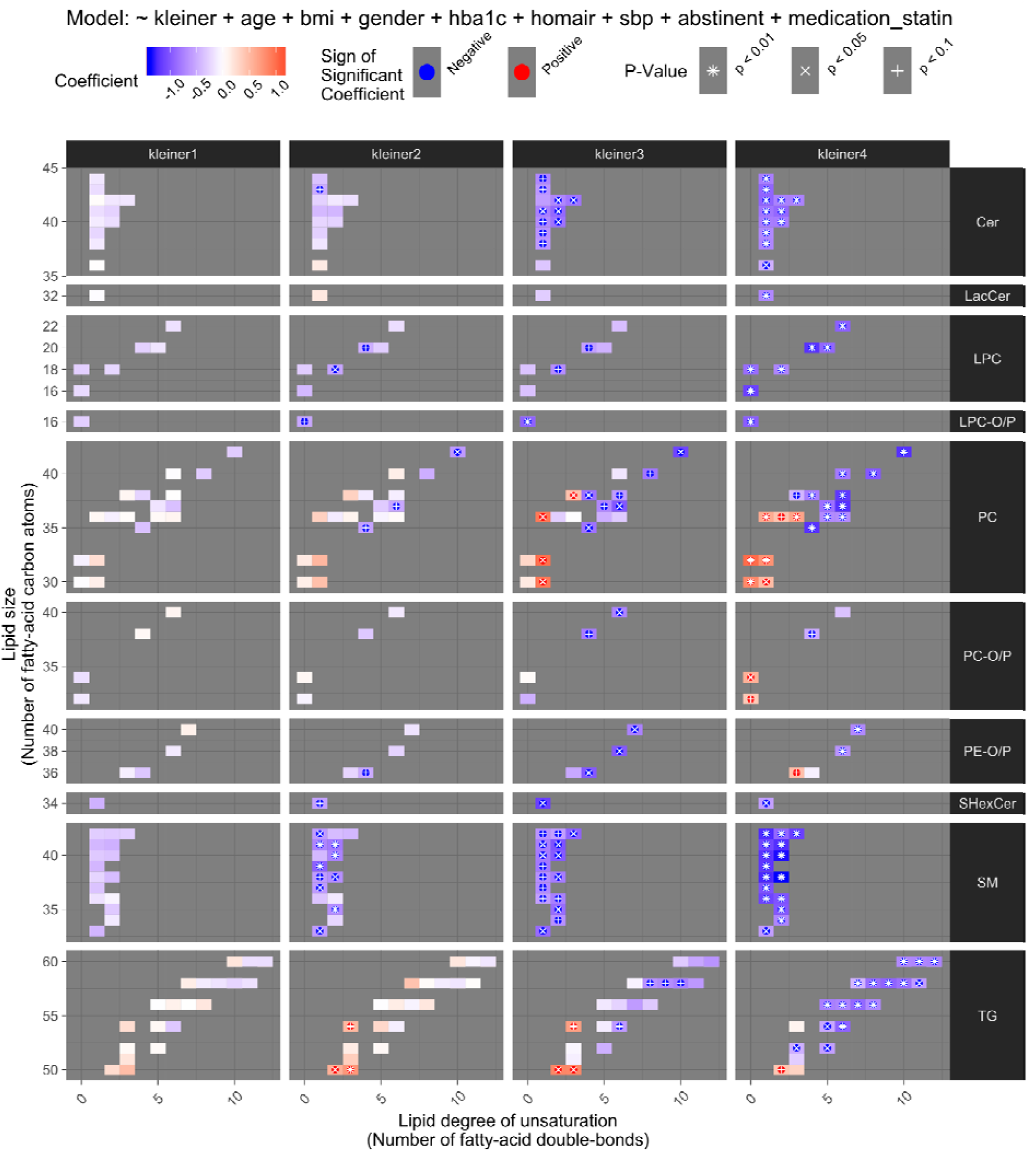
The plasma lipidome according to fibrosis stage. Plasma lipids from 10 lipid classes are affected in fibrosis. For further details, see Figure 1. **Abbreviations:** Cer, ceramides; LacCer, lactocyl-ceramides; LPC, lyso-phosphocholines; LPC-O/P, alkyl or alkenyl ether lyso-phosphocholines; PC, phosphocholines; PC-O/P, alkyl-acyl phosphocholines; PE-O/P, alkyl or alkenyl ether phosphatidylethanolamines; SHexCer, sulfatide hexocyl-ceramides; SM, sphingomyelins; TG, triacylglycerols.

In PCs and PC-O/Ps we observed a disbalance between saturated and unsaturated lipids with increasingly strong negative associations for longer, polyunsaturated lipids, contrasted by positive associations for the small, saturated lipids. We observed the same pattern in TGs.

### Inflammation-associated changes in the liver and plasma lipidomes

Liver inflammation associated with widespread lipidomic changes both in the liver and in plasma (Supplementary Figures S2 and S3). In the liver, PCs and mid-sized, monounsaturated SMs decreased progressively from mild to severe inflammation. A large number of unsaturated TGs, primarily mid-sized with less than five fatty-acid double bonds, were elevated already at minimal inflammation, with increasing numbers of TGs elevated at higher grades of inflammation. DGs, LacCers and PC-O/Ps also associated positively with inflammation.

In plasma, we observed the same negative association to inflammation in mid-sized SMs as seen in the liver. In contrast to liver, few plasma PCs showed a positive correlation with inflammation and only one TG in plasma was elevated in inflammation.

### Steatosis associated changes in the liver and plasma lipidomes

Triglycerides showed the most notable association with histological steatosis score, characterized by a broad and progressive elevation of unsaturated TGs in both liver and plasma (Supplementary Figures S4 and S5). Five DGs and four PC-O/Ps were elevated in the liver, whereas six PCs were decreased. In plasma, fewer TGs and no DGs were associated with steatosis. Interestingly, we observed a reversal in the pattern of PC-O/Ps and PCs from liver to plasma: the respective up- and downregulation in the liver reversed to down- and upregulation in plasma. Finally, SMs associated negatively with steatosis score in plasma but not in the liver.

### Integrative analysis of liver histology and the lipidomes

In an integrative analysis of ALD, we found that PCs and SMs dominated the aberrations in the liver lipidome related to fibrosis stage, whereas TGs dominated in steatosis (Figure 3). In inflammatory activity, roughly half of the significant associations with the liver lipidome were PCs, SMs, and Cers, whereas the other half were TGs, rendering the lipidome of inflammation in the liver a crossover between fibrosis and steatosis. The total number of lipids associated with liver health was greater in the liver lipidome compared to plasma (Figure 3). Overall, in the liver, the lipid class-wide changes were characterized by a decrease in PCs and SMs in fibrosis and inflammatory activity, and an elevation in DGs and TGs in inflammatory activity and steatosis (Figure 4).

**Figure 3:**
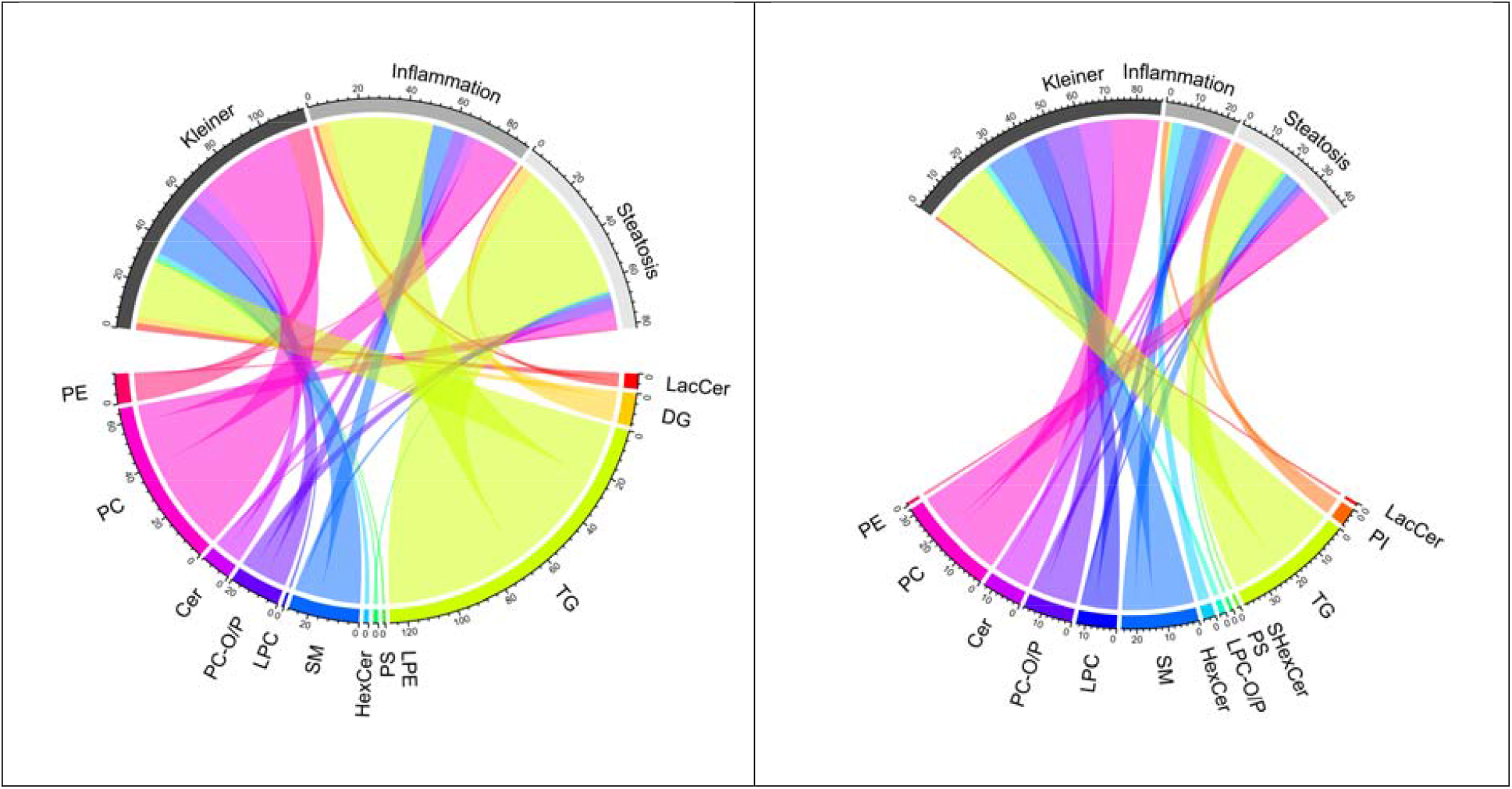
Integrative analysis of the lipidomic changes in the liver (left) and plasma (right) according to the three main types of histological changes in the liver (Kleiner fibrosis stage, inflammatory activity, and steatosis score). In a chord diagram, lipid classes are shown in distinct colors and the width of the line between a lipid class and a histological liver measure indicates the respective number of significant associations (p < 0.05) detected by the ANCOVA. Lipids are shown in the bottom-half of the circle, and liver lesions are shown in the top-half of the circle. For instance, there are approximately 60 associations between TGs in the liver and steatosis, as shown by the widest line, which is colored yellow-green in the left-side subfigure. Furthermore, large numbers of associations are observed to inflammatory activity and fibrosis. On the contrary, the number of steatosis-associated TGs in plasma is smaller, as shown by the narrower TG-Steatosis line in the right-side figure. Moreover, there are only few associations between plasma TGs and inflammatory activity, as can be seen by the almost-missing TG-Inflammation line. **Abbreviations:** ANCOVA, analysis of covariance; Cer, ceramides; DG, diacylglycerols; HexCer, hexocyl-ceramides; LacCer, lactocyl-ceramides; LPC, lyso-phosphocholines; LPC-O/P, alkyl or alkenyl ether lyso-phosphocholines; LPE, lyso-phosphatidylethanolamines; PC, phosphocholines; PC-O/P, alkyl-acyl phosphocholines; PE, phosphatidylethanolamines; PI, phosphatidylinositols; PS, phosphatidylserines; SHexCer, sulfatide hexocyl-ceramides; SM, sphingomyelins; TG, triacylglycerols.

**Figure 4:**
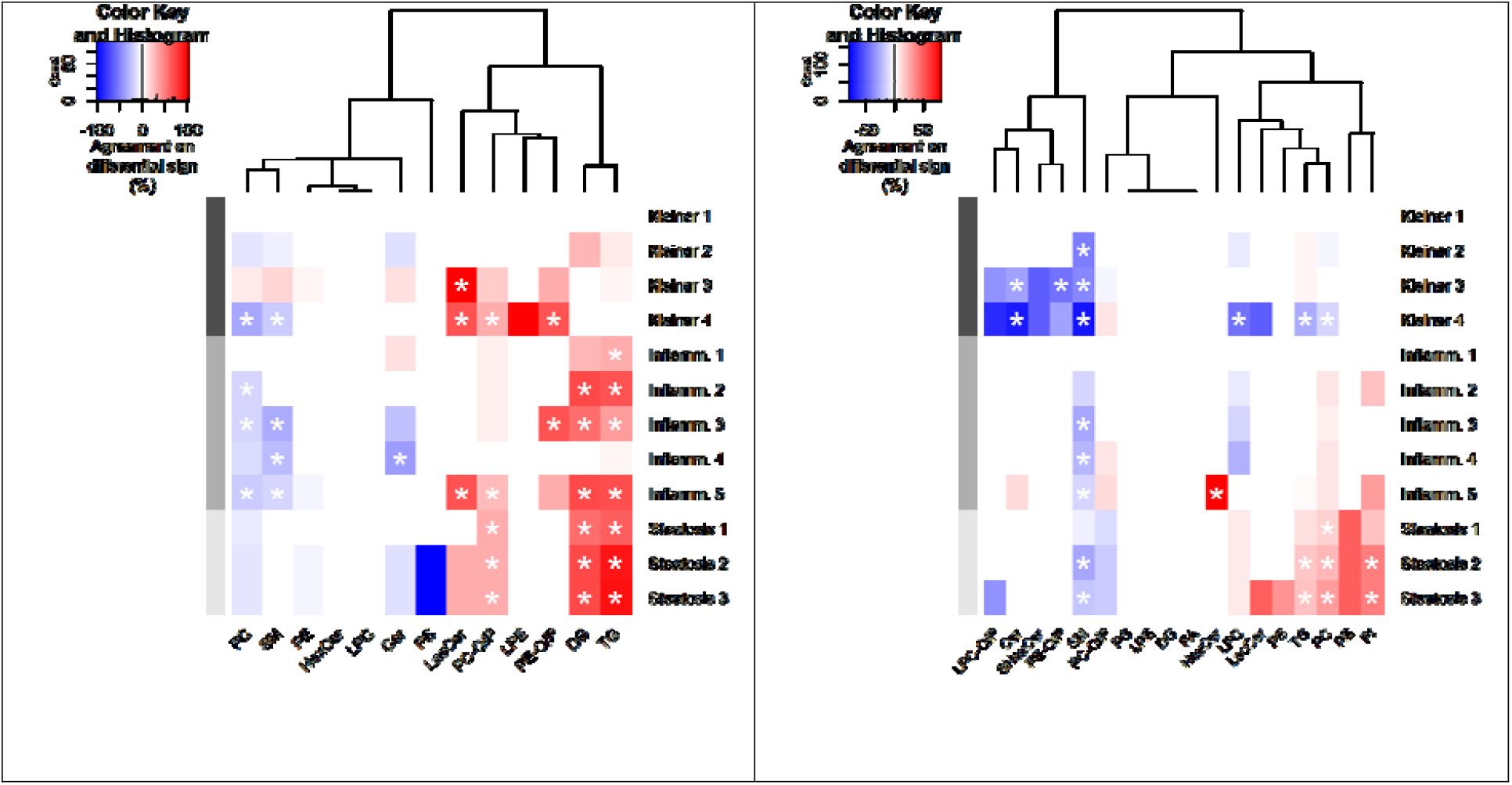
Enrichment analysis of changes in the lipidome related to liver histology, in the liver (left) and plasma (right). The enrichment is shown by lipid class (columns) and liver disease stage (rows) and an enrichment with statistical significance (p < 0.05 after correction for multiple testing) is indicated by an asterisk. The depth of the color shows the proportion of lipids with a significant association in the class, and the color shows the sign of the associations (red: positive; blue: inverse). For instance, a significant proportion of all measured SMs were decreased in cirrhosis patients (fibrosis stage F4, Kleiner 4), as indicated by the asterisk in the blue square in the “SM”-column and “Kleiner 4” row of the left-side subfigure. **Abbreviations:** Cer, ceramides; DG, diacylglycerols; FA, free fatty acids; HexCer, hexocyl-ceramides; LacCer, lactocyl-ceramides; LPC, lyso-phosphocholines; LPC-O/P, alkyl or alkenyl ether lyso-phosphocholines; LPE, lyso-phosphatidylethanolamines; PC, phosphocholines; PC-O/P, alkyl-acyl phosphocholines; PE, phosphatidylethanolamines; PG, phosphatidylglycerols; PI, phosphatidylinositols; PS, phosphatidylserines; SHexCer, sulfatide hexocyl-ceramides; SM, sphingomyelins; TG, triacylglycerols.

A total of 34, three and 11 lipid species, respectively, were associated with fibrosis, inflammatory activity or steatosis, both, in the liver and in plasma (Supplementary Figure S6). Among these liver-and-blood-associated lipids of ALD, Cers, SMs, PCs and TGs were significantly enriched in fibrosis, whereas TGs were enriched in steatosis. The most strongly associated lipids both in the liver and plasma at an early stage of ALD were the saturated sphingomyelin SM(d41:0) in fibrosis, the SM(d40:0) in hepatic inflammatory activity, and the saturated triacylglycerol TG(52:0) in steatosis (Figure 5).

**Figure 5:**
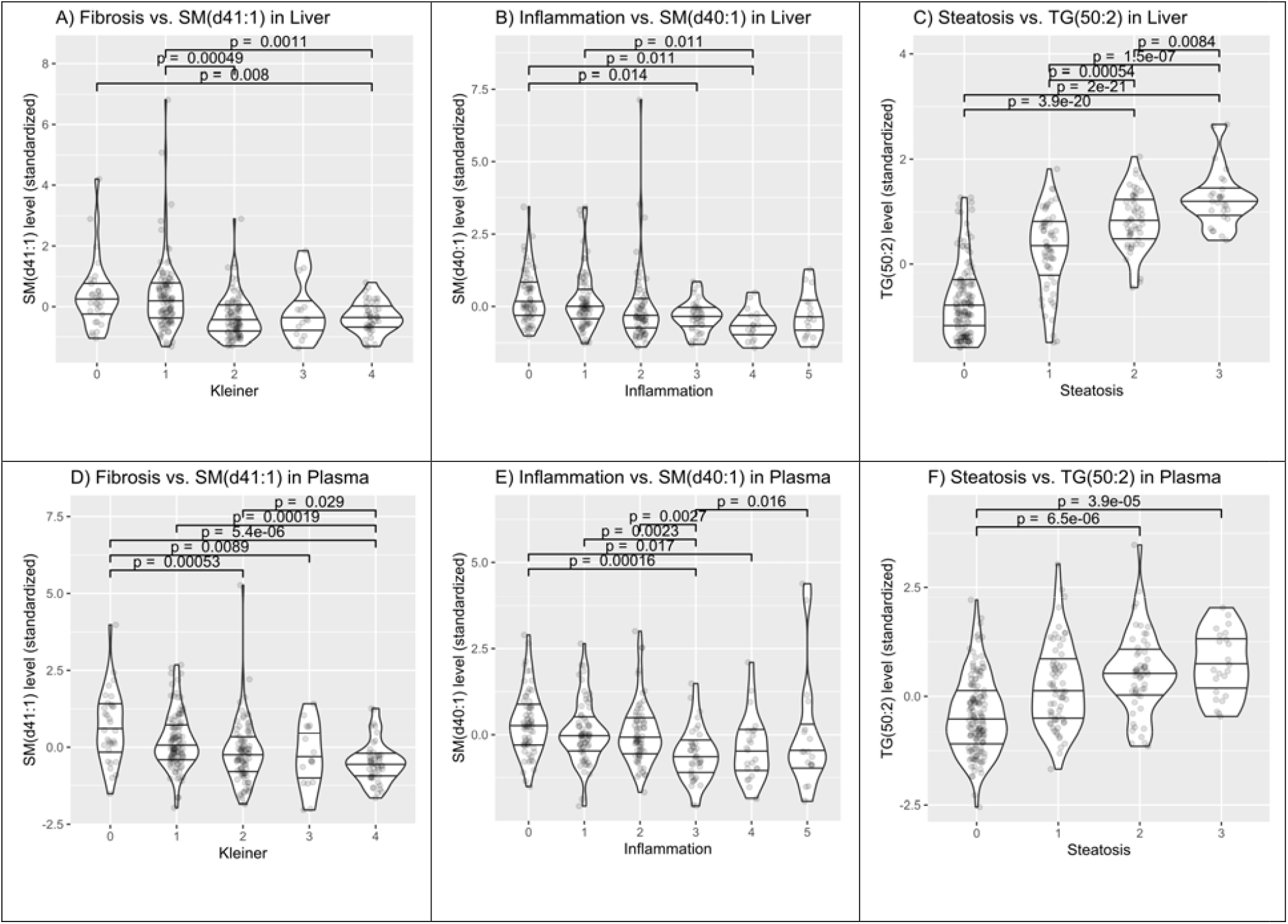
Individual lipids simultaneously aberrated, both, in the liver and in plasma. Violin plots of the levels of three individual lipid species are shown in the liver (top) and in plasma (bottom). The sphingomyelin SM(d41:1), the sphingomyelin SM(d40:1) and the triacylglycerol TG(50:2), respectively, were associated with fibrosis (left), inflammatory activity (center) and steatosis (right), simultaneously in the liver and in plasma.

### Sphingolipid-regulation in alcohol-related fibrosis

Sphingolipid dysregulation in ALD is a novel finding, which lead us to explore its mechanisms further. We therefore aimed to validate our findings *post-hoc* by investigating gene regulation and expression of the sphingolipid biosynthesis and metabolism pathways. An integrative combinatorial analysis of the transcriptome using KEGG pathways and PathWalks revealed clusters of pathways, including sphingolipid metabolism. We found a total of 44 measured miRNAs potentially regulating 25 genes in the sphingolipid metabolism pathway. Three of the miRNAs, namely, miR-21-5p, miR-24-3p and mir-146a-5p, were significantly upregulated during the course of ALD progression (Supplementary Figure S7), and are targeting most of the genes.

Second, we measured mRNA transcripts in whole blood from the sphingolipid pathways (Supplementary Figures S9 and S10). We found an overweight of upregulated enzymes involved in the sphingosine-1-phosphate (S1P) synthesis and metabolic pathways, whereas *de novo* synthesis was not clearly aberrated (Supplementary Figures S11-S13).

Finally, we investigated the causality between ALD and sphingomyelin levels, and the specificity of the association to ALD, via Mendelian randomization meta-analysis in the FinnGen and UK biobanks (Supplementary Methods). We confirmed that genetic susceptibility to ALD was associated with genetic susceptibility to lower SM levels, indicating a causal relationship between ALD and a decrease in the blood SM level (single nucleotide polymorphism (SNP)-effect: -0.032; 95% CI: -0.055,-0.0093; p=0.0058; Supplementary Figures S14-S15), but not vice versa. This causal association was specific to ALD and did not exist between NAFLD or alcohol addiction and SM levels.

## DISCUSSION

In this study, we report that alcohol-related liver disease (ALD) is associated with comprehensive changes in the liver and plasma lipidomes across the full spectrum of histological changes in the liver. In particular, long- and mid-chained sphingomyelins exhibit a strong and progressive depletion in liver fibrosis, mirrored by a progressive depletion in hepatic inflammation. This depletion is independent of metabolic comorbidities, current alcohol intake, and use of lipid-lowering drugs. In contrast, liver steatosis is characterised by elevated acylglycerols. Our analyses of gene regulation indicate that the sphingolipid depletion is due to upregulated break-down via sphingosine-1-phosphate pathways and not due to decreased synthesis. Mendelian randomization suggests that ALD causes low sphingomyelin levels, and not vice-versa. This aberration is specific to ALD, thus, not present in NAFLD.

Lipidomics, the systematic study of the molecular lipid profiles, has thus far mostly been used to describe diseases associated with metabolic dysfunction, although studies are emerging in alcohol-related liver disease [15, 27, 28]. Lipidomics studies in rodent models of NAFLD found that elevated sphingolipids (ceramides and sphingomyelins) associated with steatosis and steatohepatitis [12, 29]. However, a human case-control study found that persons with NAFLD and steatohepatitis had significantly lower levels of Cers, in addition to PCs and PEs, than patients with simple steatosis or healthy controls. Two studies on persons undergoing bariatric surgery found increased ceramides in the livers of patients with NAFLD due to insulin resistance as well as in NASH [30, 31]. In accordance with the two latter studies, ceramide inhibition improved steatosis, attenuated insulin resistance and protected against obesity in rodent models [32-34]. Presently, we are not aware of studies investigating sphingomyelins or phospholipids in NAFLD fibrosis.

Lipidomics research in alcohol-related liver disease has mostly focused on triglycerides as they are the major lipid class accumulating in hepatocytes [10]. The scarce evidence that exists on sphingolipids in ALD contrasts with our findings: Two case-control studies found increased levels of sphingolipids in ALD patients with cirrhosis [16, 35], and a rodent study linked alcohol intake to sphingolipid accumulation [36]. Another rodent study linked upregulated ceramide gene expression to steatohepatitis and pericellular fibrosis, but found conflicting evidence with regards to actual levels of Cers [37]. A case-control study found increased activity of acid sphingomyelinase, the enzyme that hydrolyses sphingomyelin into ceramide, in patients hospitalized for alcohol detoxification [38].

Given the existing evidence, our results are unexpected, since we found markedly reduced Cers and SMs in the plasma and livers of ALD patients with increasing severity of fibrosis and hepatic inflammation. To our knowledge, only one other recent study has reported ceramide depletion with severe fibrosis: The circulating levels of Cers were reduced in 12 persons with cirrhosis, out of a total of 75 persons with chronic hepatitis C virus (HCV) [39]. Our finding of progressively elevated TGs and DGs with liver steatosis severity is confirmed by existing evidence, both, in NAFLD and ALD [10, 14].

Sphingolipids are not merely proinflammatory or lipotoxic. Rather, Cers, SMs and their end-product sphingosine-1-phosphate (S1P) are readily convertible via the same enzymatic pathways (Figure S16), and aberrations in two points in the pathway may have opposing effects [12, 29]: whereas Cers and SMs are associated to apoptosis and growth arrest, S1P has been ascribed to proinflammatory activity as well as to cell survival, growth, and proliferation [12]. Similarly, the roles of Cers and SMs as signaling molecules on a cell’s lipid bi-layer differs from their effects in the cell nucleus or intracellular organelles (Figure S14).

Our cohort study was not designed for an in-depth exploration of the possible mechanisms of sphingolipid depletion. For example, we neither directly measured S1P nor studied the modulation of sphingolipids. Based on miRNA and circulating gene expression analyses, however, we propose that progressive ALD causes an inflammatory cascade, which increases breakdown, or utilization, of Cers and SMs, ultimately leading to their depletion.

Two drugs are known to modulate the Cer-SM-S1P pathway: Suramin inhibits the S1P receptor 3. It appears to attenuate bile duct ligation induced fibrosis, but it also promotes inflammation [40]. Fingolimod is a S1P receptor modulator. In a mouse model of NAFLD, it decreased steatosis, but increased inflammation and had no effect on fibrosis [41]. Dietary sphingolipids have been shown to diminish intestinal absorption of cholesterol, fatty acids and TGs, and reduce hepatic lipid uptake [42].

Our study suggests a response in SMs according to their length and saturation, as their depletion was not entirely class-wide in the liver. A few small, saturated and mono-unsaturated SMs were elevated in fibrosis and inflammation in the liver. To our knowledge, this has not been investigated previously, but future mass-spectrometry methods may be able to cover a wider range of lipid species and classes than what is currently possible, thereby shedding more detailed light on the role of individual lipids.

The main limitation of the present study is the cross-sectional study design, which prohibits causal inference. We sought to complement this by investigating the regulation and expression of genes involved in the sphingolipid pathways. We found evidence of an upregulation of miRNAs involved in the activation of sphingolipid metabolism, and dysregulated transcription levels of enzymes in the same pathway. The transcriptomics approach collects mRNA from whole blood, where the main contribution of the transcriptome comes from leukocytes. The string presence of the immune system in the whole blood transcriptome may cover other relevant pathways. Part of this circulating transcriptome, however, has been demonstrated to arise from tissues (see, e.g., [43][44] [45]). Thus, aberrations of the liver and related systems could be reflected not only in the lipidome but also in the transcriptome of the blood.

Advantages of this study are the relatively large number of deep-phenotyped participants, the careful adjustment to potential confounders, the interlaboratory-validated lipidomics method, and the causal validation by Mendelian randomization in independent cohorts.

In conclusion, global lipidomics analyses reveal that alcohol-related liver fibrosis is associated with a progressive depletion of sphingomyelins, ceramides and phosphatidylcholines in the circulation and in the liver. This contrasts findings in diseases of metabolic dysfunction, indicating a dissimilarity in the pro-fibrotic and pro-inflammatory mechanisms between non-alcoholic and alcohol-related liver diseases.

## Data Availability

The data are available by contact to, after approval from the Danish Data Protection Agency.

## Abbreviations

ALD: alcohol-related liver disease
ANCOVA: analysis of covariance
BMI: body mass index
Cer: ceramides
DG: diacylglycerols
FA: free fatty acids
HbA1C: glycated hemoglobin
HexCer: hexocyl-ceramides
HOMA-IR: homeostasis model assessment for insulin resistance
LacCer: lactocyl-ceramides
LPC: lyso-phosphocholines
LPC-O/P: alkyl or alkenyl ether lyso-phosphocholines
LPE: lyso-phosphatidylethanolamines
NAFLD: non-alcoholic fatty liver disease
PC: phosphocholines
PC-O/P: alkyl-acyl phosphocholines
PE: phosphatidylethanolamines
PE-O/P: alkyl or alkenyl ether phosphatidylethanolamines
PG: phosphatidylglycerols
PI: phosphatidylinositols
PS: phosphatidylserines
S1P: sphingosine-1-phosphate
SHexCer: sulfatide hexocyl-ceramides
SM: sphingomyelins
TG: triacylglycerols

## ACKNOWLEDGEMENTS

We wish to thank Louise Skovborg Just and Lise Ryborg, respectively, for managing the GALAXY and MicrobLiver consortia; Julie Hansen and the rest of the personnel at Center for Liver Research for assistance in the conduct of clinical investigations; Karsten Lauridsen and Louise Vestring, Heads of Department for Gastroenterology and Hepatology at Odense University Hospital; the personnel and management at Odense Municipality Alcohol Rehabilitation Centre, and OPEN Patient data Exploratory Network for data management support.

