## Supplementary Material for "Association of the Liver and Plasma Lipidomes with the Histological Stage of Alcohol-Related Liver Disease"

### SUPPLEMENTARY METHODS

#### Transcriptomics and microRNA

We measured circulating gene expression from whole blood stabilized in PAXgene® Blood RNA tubes, purified using QIAsymphony SP and PAXgene Blood RNA Kitfull blood vials. In total, we sequenced 362 samples, but excluded four samples after quality control. The transcriptome reads were mapped to human genome (GRCh38.90) using STAR (2.5.3a), and expressed genes were quantified using HTseq (0.7.2). Genes with at least two counts per million (CPM) in at least 10 individuals were included in the downstream analysis and others were excluded.

We measured the expression of 96 plasma miRNAs by semi-quantitative real-time qPCR and TaqMan MicroRNA assays. We explored further the 54 miRNAs that were expressed in >75% of the specimens. We investigated their experimentally verified gene targets through Tarbase v8.

We detected differentially expressed genes (DEGs) between ALD and healthy control using the edgeR package (3.26.4) in R with a false discovery rate of 0.05.

In an integrative analysis of the transcriptome and the Kyoto Encyclopedia of Genes and Genomes (KEGG), we inferred clusters of pathways involved in ALD with PathWalks (Karatzas, 2020, https://doi.org/10.1093/bioinformatics/btaa291 ). We used transcripts involved in the sphingolipid metabolism pathway to identify sphingolipid-regulating miRNAs.

#### Lipidomic Analysis Methodology

##### Overview

Lipidomics is normally a high-resolution mass spectrometry method focusing on detection of heavy lipids, which are present in the chloroform or methyl tert-butyl ether extract of plasma or tissue.[18] Coupled liquid chromatograph can separate molecules based of their polarity. Additional separation increases number and reliability of identified lipids. Measured in both polarities it can detect a substantial amount of lipid groups.[19] Metabolites can be further classified based on number of carbon atoms and double bonds. Most abundant groups are triacyl and diacyl glycerols (DG, TG), glycerophosphocholines including phosphocholines (PC), phosphoethanol amines (PE), sphingolipids including sphingomyelins (SM) and ceramides (CE), sterol lipids and fatty acids (FA). Lipidomics has been proven efficient to describe changes happening during metabolic syndrome, type 1 and 2 diabetes, cardiovascular diseases, non-alcoholic fatty liver disease, and alcohol-related liver disease.

**Plasma Lipidomics**

Preparation of plasma samples, instrumental analysis and data pre-processing was done according to the standard method used at Steno Diabetes Center Copenhagen. It has been described in previous works (see, e.g., Tofte, et al., 2019). The novel lipidomics analysis of liver tissue is explained here next.

**Liver Tissue Lipidomics**

Preparation of needle biopsies is based on prior methods, modified to suit the experimental design and analytical instruments. We snap-froze in liquid nitrogen approximately 5 mm of liver tissue immediately after transcutaneous liver biopsy. At Steno Diabetes Centre Copenhagen, each tissue sample were transferred from -80°C into Covaris TT05 tissue tubes, immerged into liquid nitrogen, and crushed with Covaris CryoPrep impactor CP02. Two miligrams of the powdered liver tissue were used for extraction. Exact mass was measured and written down for post-processing and normalization. 200 ul of 0.9 M NaCl in water was used to create a suspension which was vortex-mixed. Protein precipitation and extraction was made with 376 µl of chloroform : methanol (2:1, v/v) solution and 104 µL of standard mix (10 µg mL-1) solution composed of the same internal standards as in the plasma preparation procedure. After 30 min protein precipitation, the extract was centrifuged for 3 min with 10000 rpm at 4°C. 60 µl of chloroform phase was transferred into vial where it was diluted with 60 µl of clean chloroform : methanol (2:1, v/v) solution.

For quality control, extracts were also pooled together and (insert the exact amount) of the pooled samples were inserted into the chromatographic sequence. Sample preparation variability was controlled using mouse liver tissue. Instrumental analysis was the same as for plasma samples, described above.

#### Data Analysis

All reported statistical tests were corrected for multiple testing over the lipidome using the Benjamini-Hochberg method. All data analyses were done with R (version 3.6.2).

**Statistical Associations**

Associations between the lipidome and the three liver health measures – fibrosis, inflammation and steatosis – were analyzed in six distinct lines of lipid species-wise analyses: associations between the liver lipidome and fibrosis, inflammation and steatosis as well as associations between the plasma lipidome and the same three liver health measures. Statistical analysis and consecutive visualization of the results were completed in an integrated pipeline using the lipidomeR package (0.1.2; Suvitaival & Legido-Quigley, 2020), where each lipid-species was tested for associations with one of the three liver health measures at a time by analysis of covariance (ANCOVA) coupled to pairwise post-hoc tests.

In the final step of the pipeline, the result statistics were visualized in six lipidome-wide heatmaps, one for each pair of the lipidome source (liver or plasma) and liver health measure (fibrosis, inflammation and steatosis).

For instance, associations between the lipidome in the liver and fibrosis were detected with the lipidomeR pipeline as follows: (1) The measured level of a lipid species were modeled in a regression model by the categorical fibrosis variable and the continuous clinical adjustment variables. (2) The ANCOVA step detected, via the F-test, whether the measured lipid level had variation between the stages of fibrosis. Lipid species with significant variation were investigated further. (3) Pairwise post-hoc tests via the same regression model were applied to find out, which of the fibrosis stages differ from a healthy liver in terms of the lipid level. (4) Fibrosis stages were visualized as a lipidome-wide heatmap, which are detailed in the next subsection. The same procedure was repeated for the liver lipidome in association to inflammation and steatosis as well as for the plasma lipidome in association to each of the three liver health measures.

**Lipidome-Wide Heatmaps of Changes**

Associations between the lipidome and each of the three liver health measures were visualized with lipidome-wide heatmaps produced with the lipidomeR package. Two heatmaps were created for each of the three liver health measures: one for the liver lipidome and one for the plasma lipidome.

In a heatmap, the stages are shown as columns and lipids are grouped into classes that are shown as rows. In each stage-and-class panel, individual lipid species are organized by size (y-axis) and degree of unsaturation (x-axis). Each colored rectangle in a panel corresponds to one lipid species and the color of the rectangle shows the difference in the level of the lipid species between the fibrosis stage in question (column) and a healthy liver.

Associations with statistical significance after correction for multiple testing are highlighted with asterisk, cross and plus signs, respectively, for $p < 0.01$, 0.05 and 0.1.

**Integrative Analysis of Associations Between Liver Health and the Lipidome**

Overview of the associations between the lipidome and the three liver health measures were visualized as chord diagrams using the circlize package (0.4.13 Gu, *et al.*, 2014). One diagram was created for the liver lipidome and one for the plasma lipidome. In a chord diagram, lipid classes are shown in distinct colors and the width of the line between a lipid class and a liver health measure indicates the number of significant associations ($p < 0.05$) detected by the ANCOVA.

**Enrichment Analysis of Aberrations in Specific Stages of Liver Disease**

The detected associations were further interpreted in the context of lipid pathways by conducting an enrichment analysis: Based on the results of the post-hoc comparisons (see the Subsection "Statistical Associations"), lipid classes with more associations with statistical significance than expected were identified for each liver health measure at each of its stages. Again, two distinct lines of enrichment analyses were done: one for the liver lipidome and one for the plasma lipidome.

The consensus of aberrations in each lipid class and post-hoc comparison pair were visualized as a heatmap using the gplots package (3.1.1). In the heatmap, the liver health stages and lipid classes are shown as rows and columns, respectively. The depth of the color shows the proportion of lipids with a significant association in the class, and the color shows the sign of the associations (red: positive; blue: inverse). The columns were re-ordered with hierarchical clustering to aid identification and interpretation of lipidomic patterns. For each lipid class and post-hoc comparison pair, enrichment was tested with the binomial test. Significant enrichments were highlighted with an asterisk.

#### Causality between Liver Disease and Circulating Sphingomyelin via Mendelian Randomization

We used Mendelian randomization meta-analysis with the inverse-variance weighted estimator in large general population cohorts to externally validate the relation between ALD and circulating SM levels. We addressed three questions: First, does ALD cause a change in the SM level? Second, does NAFLD cause a change in the SM level? Third, does alcohol addiction cause a change in the SM level?

Genetic risk factors specifically to ALD or NAFLD instead of liver disease in general were publicly available in the FinnGen Biobank (R2). We used 13 single-nucleotide polymorphisms (SNPs) associated with ALD at p<10^-5 as instrumental variables for ALD in 539 persons with ALD and 95,960 controls. Similarly, 10 SNPs were available as instruments for NAFLD in 272 persons with NAFLD and 96,227 controls. Genetic risk-factors to alcohol addiction were available in the UK Biobank with 29 SNPs associated at p<10^-5 (Neale Lab, R2; Bycroft, et al., 2018) in 2778 persons ever addicted to alcohol and 3736 controls. SM level from a multinational European pan-cohort of 13,476 participants (Kettunen, et al., 2016) was used as the outcome in all three MR analyses.

### SUPPLEMENTARY RESULTS

#### Identification of Lipids

##### List of Identified Lipids in the Liver

**Supplementary Table S1: List of lipids detected and identified in the liver samples.** Name of the lipid class according to LIPID MAPS.

| Glycerolipids | | Glycerolipids | Sphingolipids | Sphingolipids |
|---|---|---|---|---|
| **Triacyl-glycerols** | | **Diacyl-glycerols** | **Phospho-sphingolipids** | **Ceramides** |
| TG(40:1) | TG(52:6) | DG(32:0) | SM(30:1) | Cer(34:1) |
| TG(42:0) | TG(52:7) | DG(34:0) | SM(32:0) | Cer(40:1) |
| TG(42:1) | TG(53:2) | DG(34:1) | SM(32:1) | Cer(40:2) |
| TG(42:2) | TG(53:4) | DG(34:2) | SM(34:0) | Cer(41:1) |
| TG(42:3) | TG(53:5) | DG(36:0) | SM(34:1) | Cer(41:2) |
| TG(44:0) | TG(54:1) | DG(36:2) | SM(36:1) | Cer(42:1) |
| TG(44:1) | TG(54:3) | DG(36:3) | SM(36:2) | Cer(42:3) |
| TG(44:2) | TG(54:4) | | SM(38:0) | HexCer(41:1) |
| TG(45:0) | TG(54:5) | | SM(38:1) | HexCer(42:1) |
| TG(46:3) | TG(54:7) | | SM(39:1) | LacCer(34:1) |
| TG(46:5) | TG(54:8) | | SM(40:0) | LacCer(42:2) |
| TG(48:1) | TG(55:1) | | SM(40:1) | |
| TG(48:3) | TG(55:2) | | SM(41:1) | |
| TG(48:4) | TG(55:3) | | SM(42:1) | |
| TG(48:5) | TG(55:4) | | SM(42:2) | |
| TG(49:2) | TG(56:1) | | SM(43:1) | |
| TG(49:3) | TG(56:2) | | SM(43:2) | |
| TG(50:1) | TG(56:3) | | SM(44:2) | |
| TG(50:2) | TG(56:4) | | | |
| TG(50:3) | TG(56:5) | | | |
| TG(50:5) | TG(58:2) | | | |
| TG(50:6) | TG(58:3) | | | |
| TG(51:2) | TG(58:4) | | | |
| TG(51:3) | TG(58:5) | | | |
| TG(52:2) | TG(58:7) | | | |
| TG(52:3) | TG(60:3) | | | |
| TG(52:4) | TG(60:9) | | | |

**Supplementary Table S1 (Continued)**

| **Glycero-phospholipids** | | **Glycero-phospholipids** | **Glycero-phospholipids** |
|---|---|---|---|
| **Glycerophospho-cholines** | | **Glycerophospho-ethanolamines** | **Glycerophospho-serines** |
| PC(30:0) | PC(40:4) | PE(34:1) | PS(38:6) |
| PC(30:1) | PC(40:5) | PE(34:2) | |
| PC(31:1) | PC(40:6) | PE(36:1) | |
| PC(32:0) | PC(40:7) | PE(36:2) | |
| PC(32:1) | PC(40:8) | PE(36:3) | |
| PC(32:2) | PC(42:10) | PE(36:4) | |
| PC(33:0) | PC-O/P(32:0) | PE(36:5) | |
| PC(33:1) | PC-O/P(34:0) | PE(38:1) | |
| PC(34:1) | PC-O/P(34:1) | PE(38:3) | |
| PC(34:2) | PC-O/P(36:1) | PE(38:4) | |
| PC(34:3) | PC-O/P(36:2) | PE(38:5) | |
| PC(34:4) | PC-O/P(36:4) | PE(38:6) | |
| PC(35:1) | PC-O/P(38:6) | PE(40:5) | |
| PC(35:4) | PC-O/P(40:3) | PE(40:6) | |
| PC(36:1) | PC-O/P(40:6) | PE-O/P(34:2) | |
| PC(36:2) | PC-O/P(42:2) | PE-O/P(38:5) | |
| PC(36:3) | PC-O/P(42:4) | PE-O/P(40:7) | |
| PC(36:4) | PC-O/P(42:5) | LPE(20:3) | |
| PC(36:5) | LPC(18:1) | | |
| PC(36:6) | | | |
| PC(37:2) | | | |
| PC(37:4) | | | |
| PC(38:2) | | | |
| PC(38:3) | | | |
| PC(38:4) | | | |
| PC(38:5) | | | |
| PC(38:6) | | | |

#### List of Identified Lipids in Plasma

**Supplementary Table S2: List of lipids detected and identified in plasma samples.** Name of the lipid class according to LIPID MAPS.

| **Glycerolipids** | | | **Glycerolipids** | **Sphingolipids** |
|---|---|---|---|---|
| **Triacyl-glycerols** | | | **Diacyl-glycerols** | **Phospho-sphingolipids** |
| TG(42:0) | TG(52:7) | TG(58:9) | DG(36:1) | SM(30:1) |
| TG(42:1) | TG(53:1) | TG(60:10) | DG(36:2) | SM(31:1) |
| TG(44:0) | TG(53:3) | TG(60:11) | DG(36:3) | SM(32:1) |
| TG(44:1) | TG(53:5) | TG(60:12) | DG(38:1) | SM(32:2) |
| TG(46:0) | TG(54:0) | | DG(40:6) | SM(33:1) |
| TG(46:1) | TG(54:1) | | | SM(34:2) |
| TG(46:2) | TG(54:2) | | | SM(35:2) |
| TG(47:2) | TG(54:3) | | | SM(36:1) |
| TG(48:1) | TG(54:5) | | | SM(36:2) |
| TG(48:2) | TG(54:6) | | | SM(37:1) |
| TG(49:0) | TG(54:8) | | | SM(38:0) |
| TG(49:1) | TG(55:2) | | | SM(38:1) |
| TG(49:2) | TG(55:3) | | | SM(38:2) |
| TG(49:3) | TG(55:4) | | | SM(39:1) |
| TG(50:0) | TG(56:1) | | | SM(40:1) |
| TG(50:1) | TG(56:2) | | | SM(40:2) |
| TG(50:2) | TG(56:3) | | | SM(40:3) |
| TG(50:3) | TG(56:5) | | | SM(41:1) |
| TG(50:4) | TG(56:6) | | | SM(41:2) |
| TG(50:5) | TG(56:7) | | | SM(42:1) |
| TG(51:2) | TG(56:8) | | | SM(42:2) |
| TG(51:3) | TG(56:9) | | | SM(42:3) |
| TG(51:4) | TG(58:10) | | | |
| TG(52:0) | TG(58:11) | | | |
| TG(52:1) | TG(58:2) | | | |
| TG(52:3) | TG(58:7) | | | |
| TG(52:5) | TG(58:8) | | | |

**Supplementary Table S2 (Continued)**

| **Sphingolipids** | **Glycero-phospholipids** | | | **Glycero-phospholipids** |
|---|---|---|---|---|
| **Ceramides** | **Glycerophospho-cholines** | | | **Glycerophospho-ethanolamines** |
| Cer(34:1) | PC(28:0) | PC(38:6) | LPC(16:1) | PE(34:1) |
| Cer(36:1) | PC(30:0) | PC(40:4) | LPC(18:0) | PE(36:3) |
| Cer(37:1) | PC(30:1) | PC(40:5) | LPC(18:1) | PE-O/P(36:2) |
| Cer(38:1) | PC(32:0) | PC(40:6) | LPC(18:2) | PE-O/P(36:3) |
| Cer(39:1) | PC(32:1) | PC(40:8) | LPC(20:4) | PE-O/P(36:4) |
| Cer(40:1) | PC(32:2) | PC(42:10) | LPC(20:5) | PE-O/P(38:6) |
| Cer(40:2) | PC(33:0) | PC-O/P(32:0) | LPC(22:5) | PE-O/P(40:6) |
| Cer(41:1) | PC(33:2) | PC-O/P(34:0) | LPC(22:6) | PE-O/P(40:7) |
| Cer(41:2) | PC(34:1) | PC-O/P(34:1) | | LPE(16:0) |
| Cer(42:0) | PC(34:2) | PC-O/P(34:2) | | |
| Cer(42:1) | PC(34:3) | PC-O/P(36:1) | | |
| Cer(42:2) | PC(34:4) | PC-O/P(36:2) | | |
| Cer(42:3) | PC(35:1) | PC-O/P(36:3) | | |
| Cer(43:1) | PC(35:2) | PC-O/P(36:4) | | |
| Cer(43:2) | PC(35:4) | PC-O/P(38:3) | | |
| Cer(44:1) | PC(36:0) | PC-O/P(38:4) | | |
| HexCer(34:1) | PC(36:1) | PC-O/P(40:3) | | |
| HexCer(40:1) | PC(36:2) | PC-O/P(40:4) | | |
| HexCer(41:1) | PC(36:3) | PC-O/P(40:5) | | |
| HexCer(42:1) | PC(36:4) | PC-O/P(40:6) | | |
| HexCer(42:2) | PC(36:5) | PC-O/P(42:4) | | |
| LacCer(32:1) | PC(36:6) | PC-O/P(42:5) | | |
| LacCer(34:1) | PC(37:2) | PC-O/P(44:4) | | |
| SHexCer(34:1) | PC(37:5) | LPC-O/P(16:0) | | |
| SHexCer(34:2) | PC(37:6) | LPC-O/P(18:0) | | |
| | PC(38:3) | LPC(14:0) | | |
| | PC(38:4) | LPC(16:0) | | |

**Supplementary Table S2 (Continued)**

| **Glycero-phospholipids** | **Glycero-phospholipids** | **Glycero-phospholipids** | **Fatty Acyls** |
|---|---|---|---|
| **Glycerophospho-serines** | **Glycerophospho-inositols** | **Glycerophospho-glycerols** | **Fatty Acids** |
| PS(36:2) | PI(34:1) | PG(36:3) | FA(16:0) |
| PS(38:5) | PI(34:2) | | FA(18:0) |
| PS(40:6) | PI(36:1) | | FA(18:1) |
| | PI(36:2) | | FA(18:2) |
| | PI(36:4) | | FA(20:4) |
| | PI(38:3) | | FA(22:6) |
| | PI(38:4) | | |
| | PI(38:5) | | |
| | PI(40:6) | | |

#### The Lipidome

Figure S1: Difference between the plasma lipidome in healthy controls and ALD

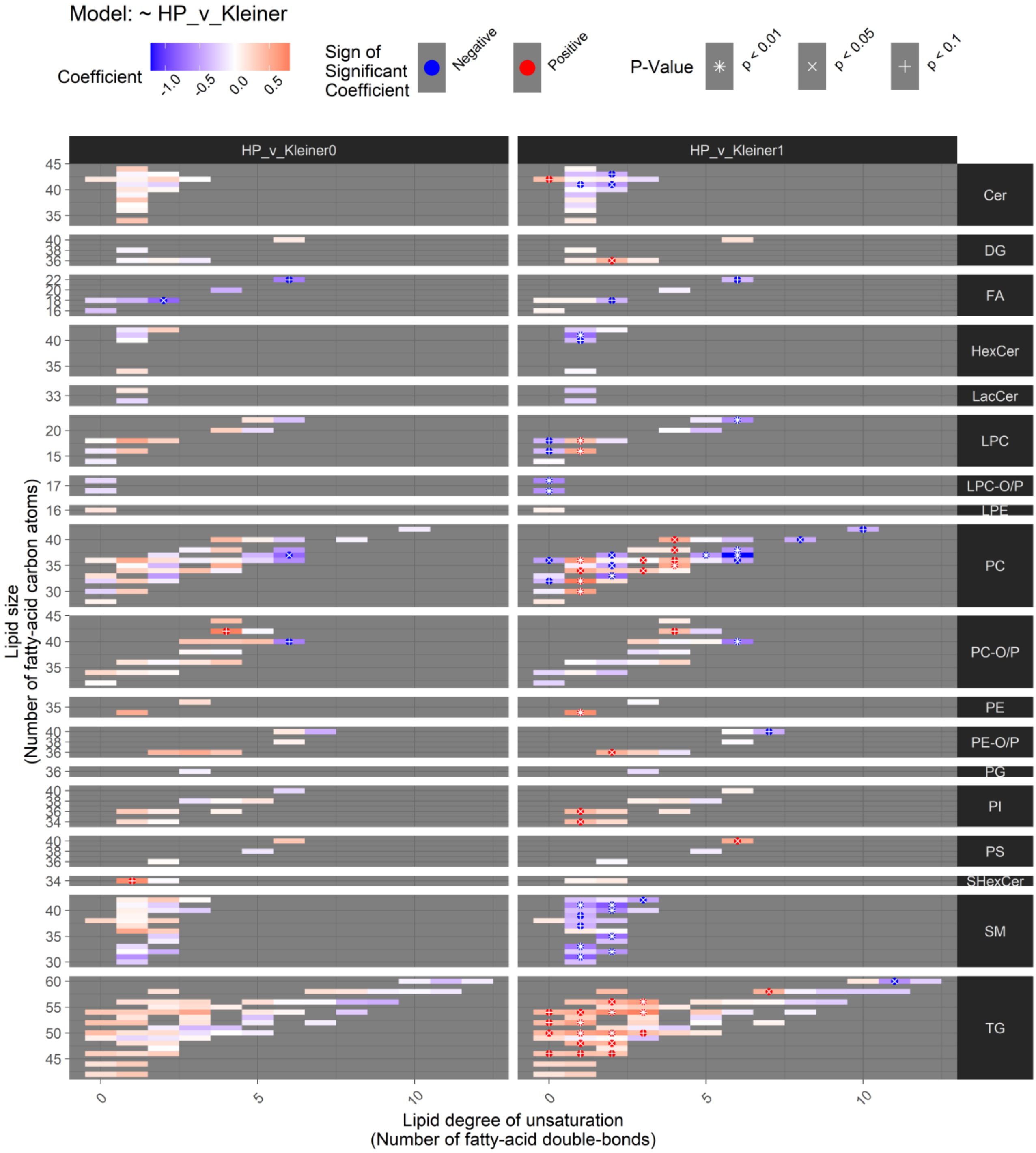

**Supplementary Figure S1: Comparison of circulating lipidomics in healthy controls (HP) versus patients with alcohol-related fibrosis stage F0 (Kleiner0) and F1 (Kleiner1).** Only the lipidome of patients with no fibrosis were comparable to matched healthy controls with significantly (P<0.05 after Benjamini Hochberg adjustment for multiple testing) lower levels of one fatty acid (FA) and one phosphocholine (PC).

#### Figure S2-S3: Inflammation-associated changes in the lipidome

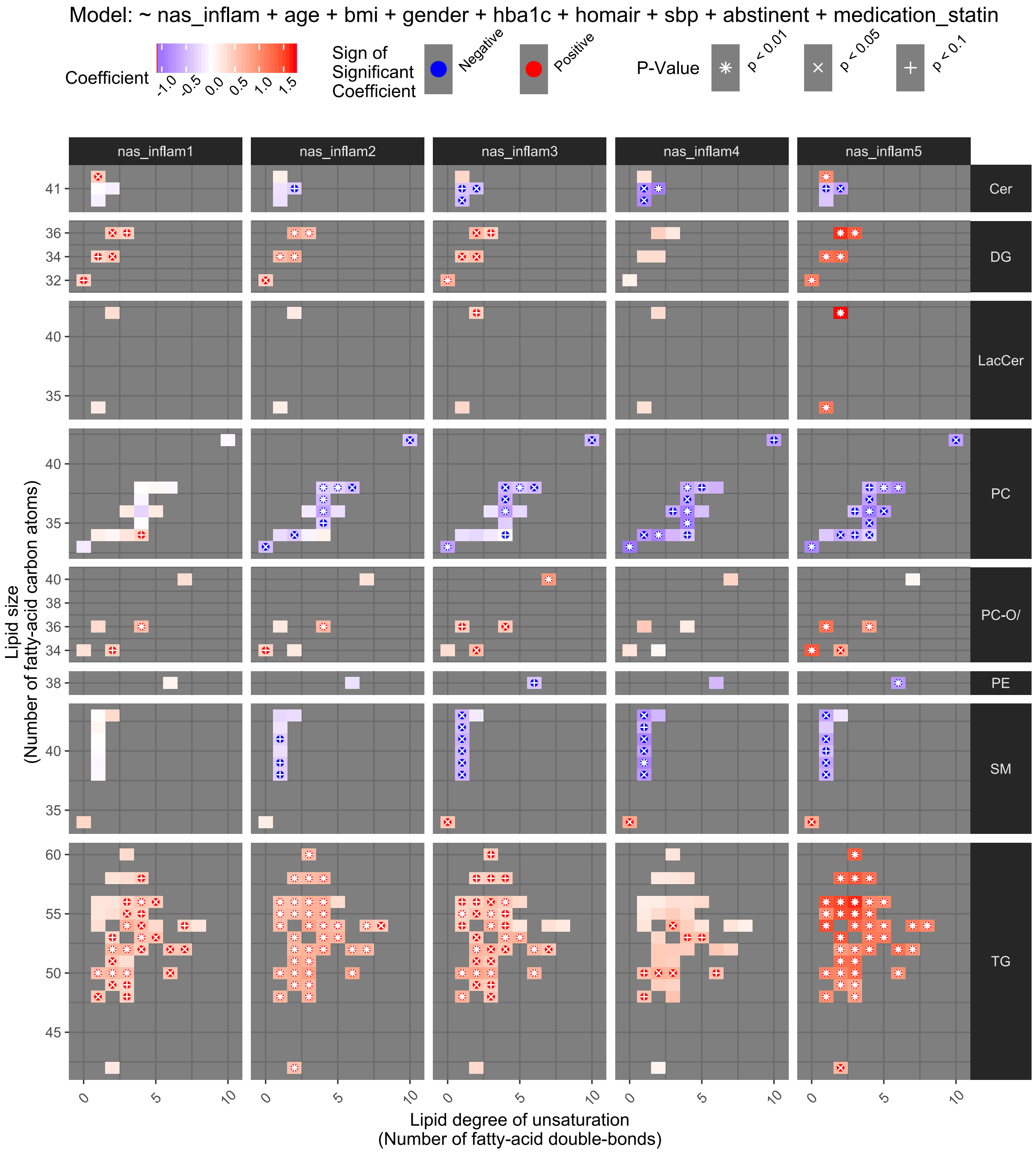

**Supplementary Figure S2: Liver lipidome according to inflammatory activity in the liver.** The associations are shown after adjustment for possible confounders (age, sex, BMI, HbA1C, HOMA-IR, systolic blood pressure, abstinence from alcohol at the time of inclusion, and statin use. Aberration in eight lipid classes (rows) across the grades of inflammation (columns, no ballooning and no lobular inflammation as reference). For each lipid class, individual lipid species are organized by size (y-axis) and degree of unsaturation (x-axis). Each colored rectangle in a panel corresponds to one lipid species and the color of the

rectangle shows the difference in the level of the lipid species between the fibrosis stage in question and a healthy liver; red is elevated, blue is reduced. Associations with statistical significance after Benjamini-Hochberg correction for multiple testing are highlighted with asterisk (*), cross (x) and plus (+) signs, respectively, for $p < 0.01$, 0.05 and 0.1.
**Abbreviations:** Cer, Ceramides; DG, diacylglycerols, LacCer, lactocyl-ceramides; PC, phosphocholines; PC-O/P, alkyl-acyl phosphocholines; PE, phosphatidylethanolamines; SM, sphingomyelins, TG, triglycerides.

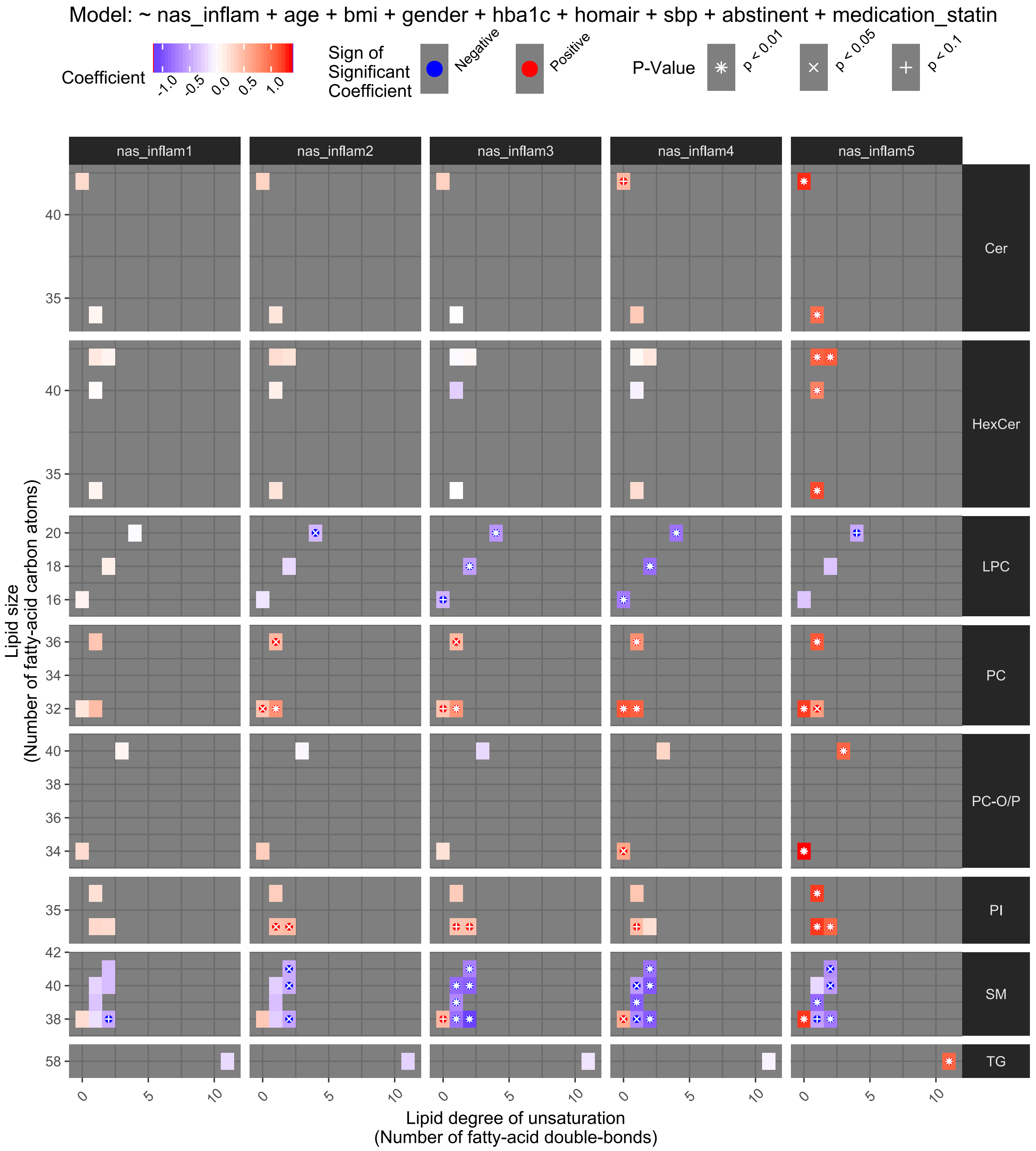

**Supplementary Figure S3: Plasma lipidome according to inflammatory activity in the liver.** Abberations are seen in eight lipid classes. See Figure S2 for further details. Abbreviations: Cer, ceramides; HexCer, hexocyl-ceramides; LPC, lyso-phosphocholines; PC, phosphocholines; PC-O/P, alkyl-acyl phosphocholines; PI, phosphatidylinositols; SM, sphingomyelins; TG, triacylglycerols.

#### Figure S4-S5: Steatosis-associated changes in the lipidome

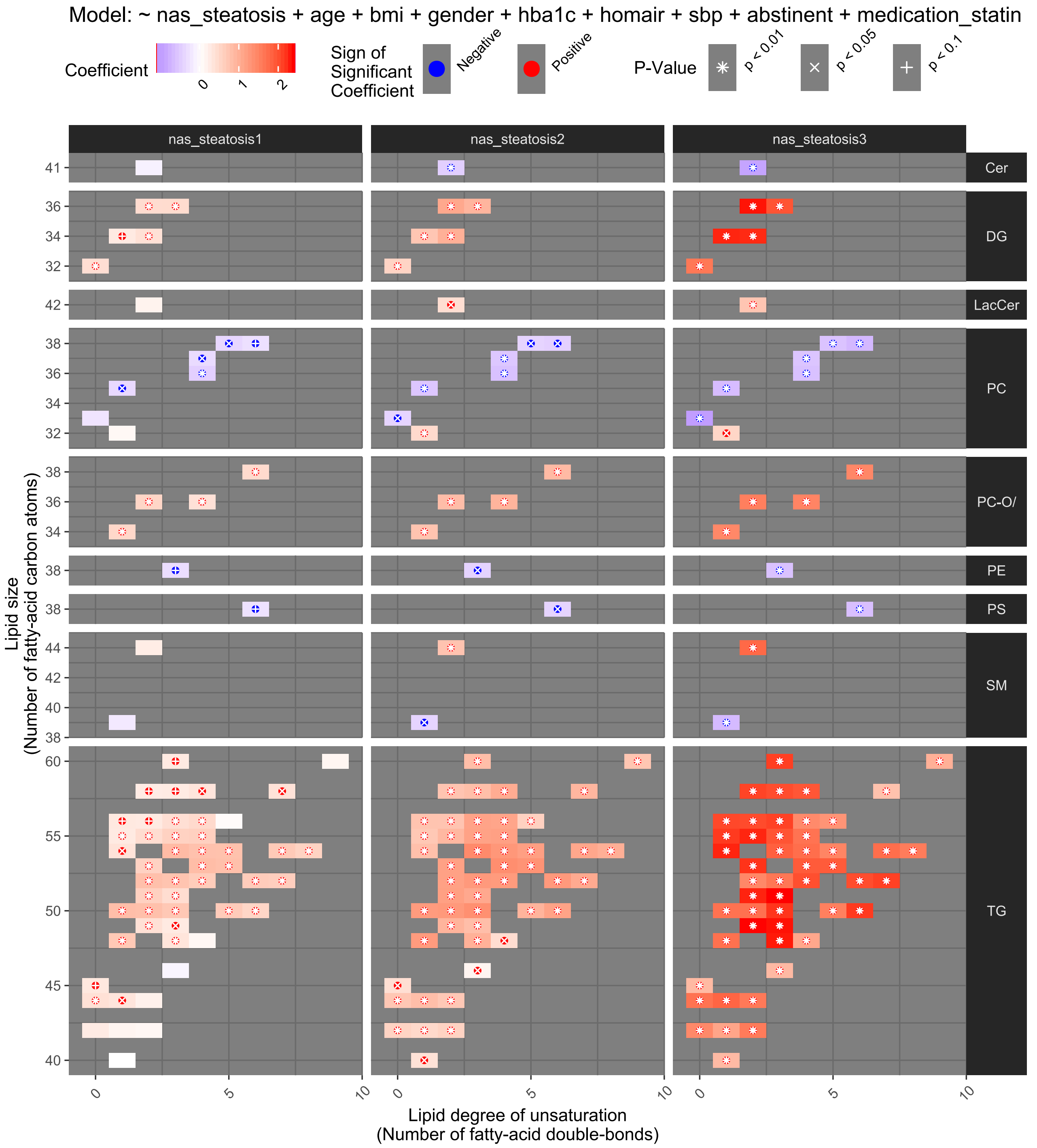

**Supplementary Figure S4: The liver lipidome according to steatosis score.** Aberations are seen in nine lipid classes. See Figure S2 for further details.

Abbreviations: Cer, ceramides; DG, diacylglycerols; LacCer, lactocyl-ceramides; PC, phosphocholines; PC-O/P, alkyl-acyl phosphocholines; PE, phosphatidylethanolamines; PS, phosphatidylserines; SM, sphingomyelins; TG, triacylglycerols.

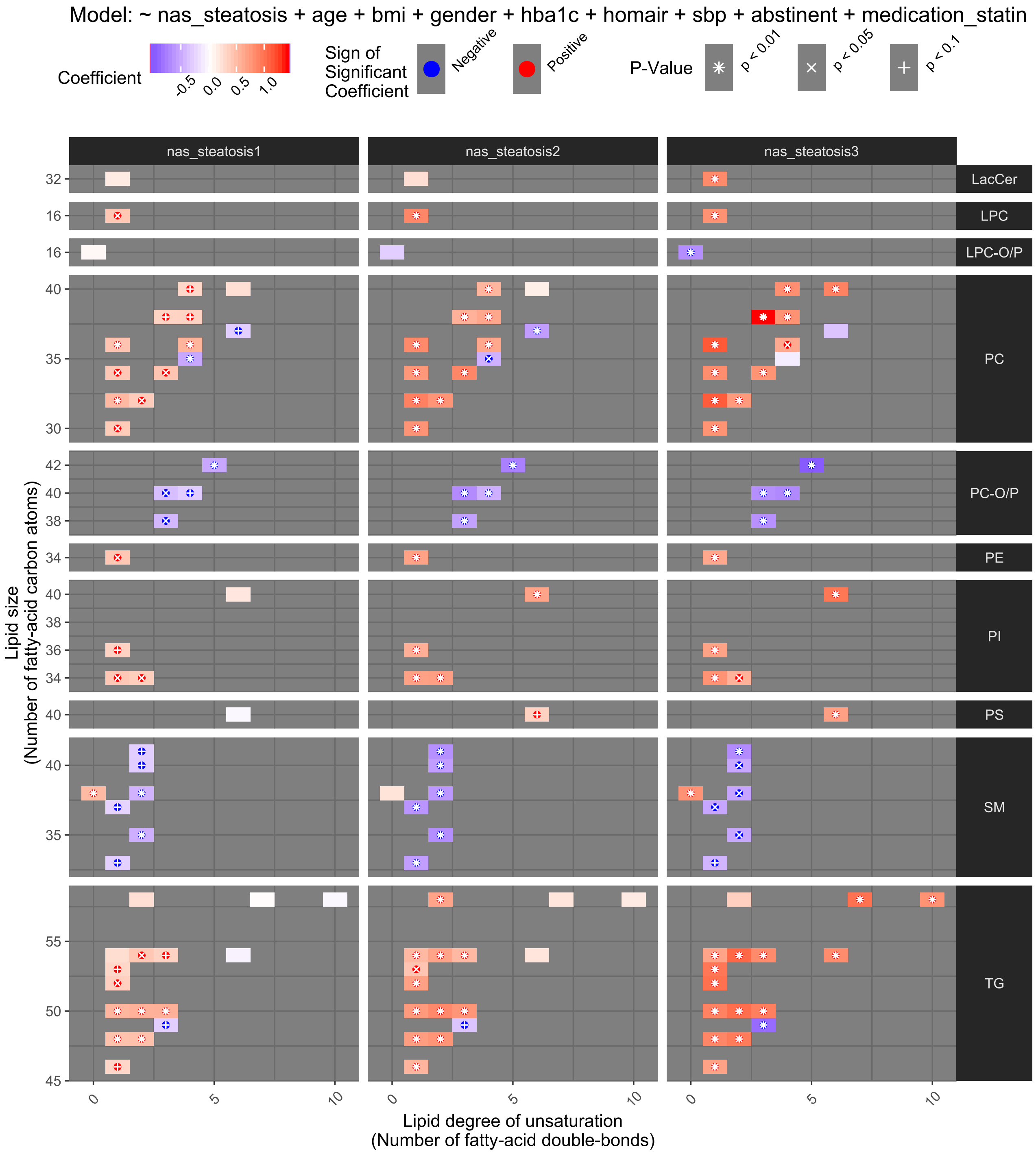

**Supplementary Figure S5: The plasma lipidome according to steatosis score.** Aberations are seen in ten lipid classes. See Figure S2 for further details.

Abbreviations: LacCer, lactocyl-ceramides; LPC, lyso-phosphocholines; LPC-O/P, alkyl or alkenyl ether lyso-phosphocholines; PC, phosphocholines; PC-O/P, alkyl-acyl phosphocholines; PE, phosphatidylethanolamines; PI, phosphatidylinositols; PS, phosphatidylserines; SM, sphingomyelins; TG, triacylglycerols.

Figure S6: Lipids simultaneously associated with alcohol-related liver disease both in the liver and plasma

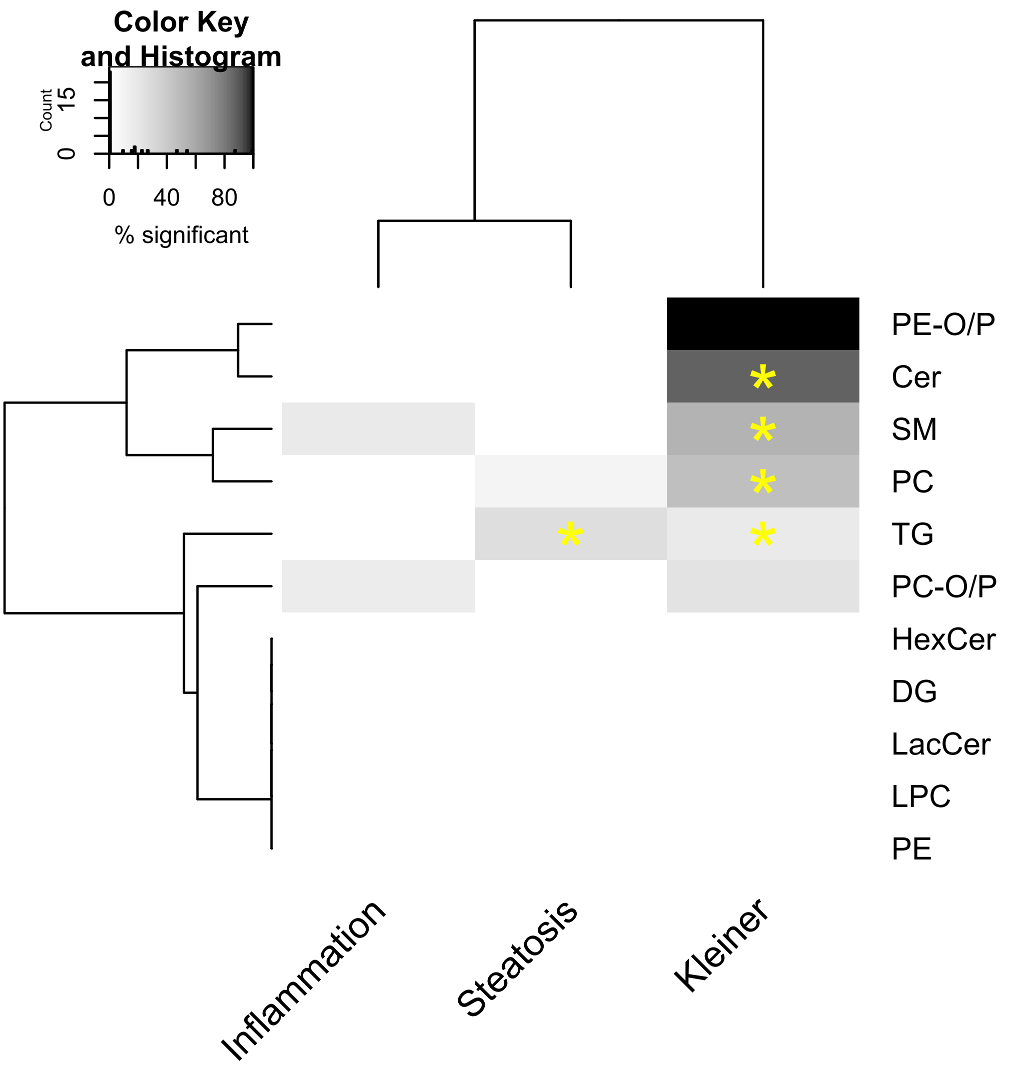

**Supplementary Figure S6: Lipid classes with an enrichment of lipids that are simultaneously associated with liver histology both in the liver and plasma.** The enrichment (proportion of lipid species within each lipid class) of these doubly-associated lipids is shown by lipid class (rows) in relation to the three measures of liver histology (fibrosis, inflammation and steatosis; columns). The proportion of lipids in the lipid class with a double association is shown by the color (white: none; black: all) and an enrichment with statistical significance is highlighted by an asterisk. The rows and columns of the heatmap are sorted by hierarchical clustering to aid interpretation.

#### MicroRNA gene regulation in the sphingolipid pathway

##### Figure S7: Circulating levels of microRNAs in alcohol-related liver disease

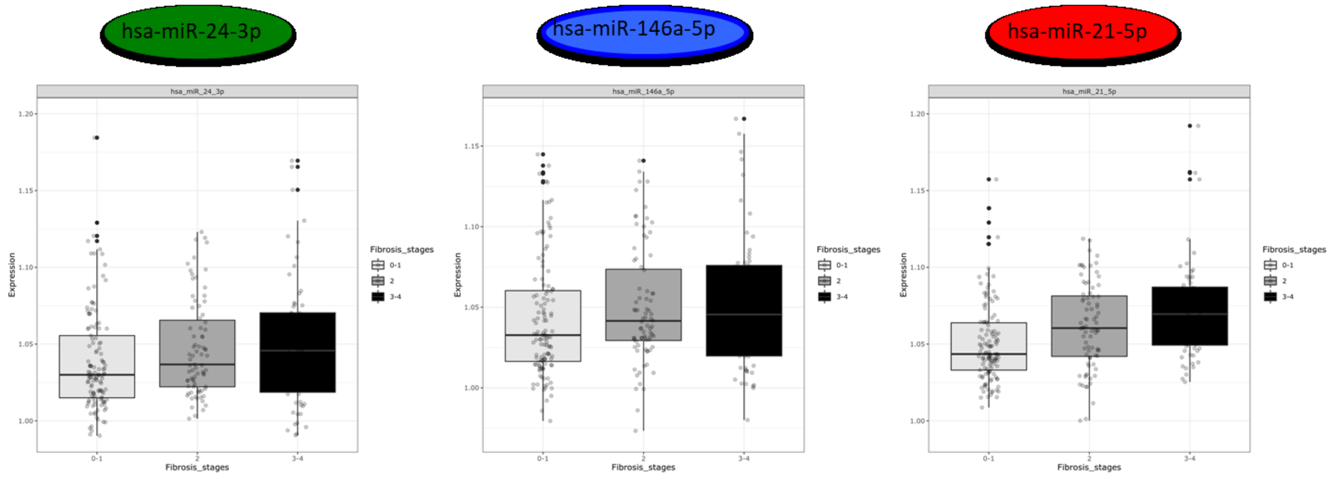

**Supplementary Figure S7: Three microRNA's involved in regulating the sphingolipid pathway had increased expression according to fibrosis severity when divided into F0-1, F2, and F3-4.** The microRNAs miR-24-3p, miR 146a-5p and miR-21-5p are shown in the three panels.

#### Figure S8: Regulated genes in the sphingolipid pathway

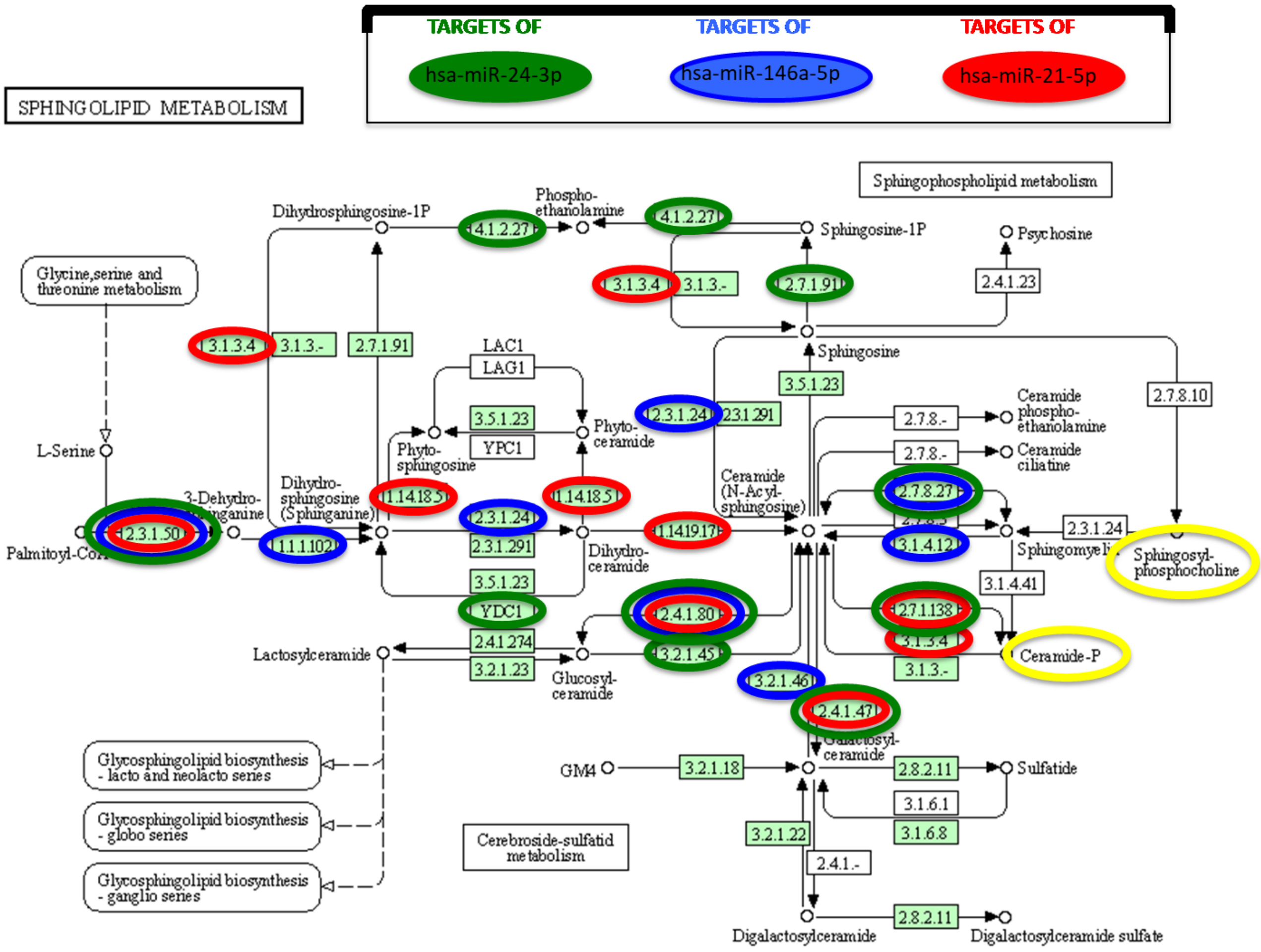

**Supplementary Figure S8: Targets of three ALD-affected microRNAs in the sphingolipid metabolism pathway.**

#### Circulating transcriptome of regulated genes in the sphingolipid pathways

Figure S9-S10: Overview of the sphingolipid pathways

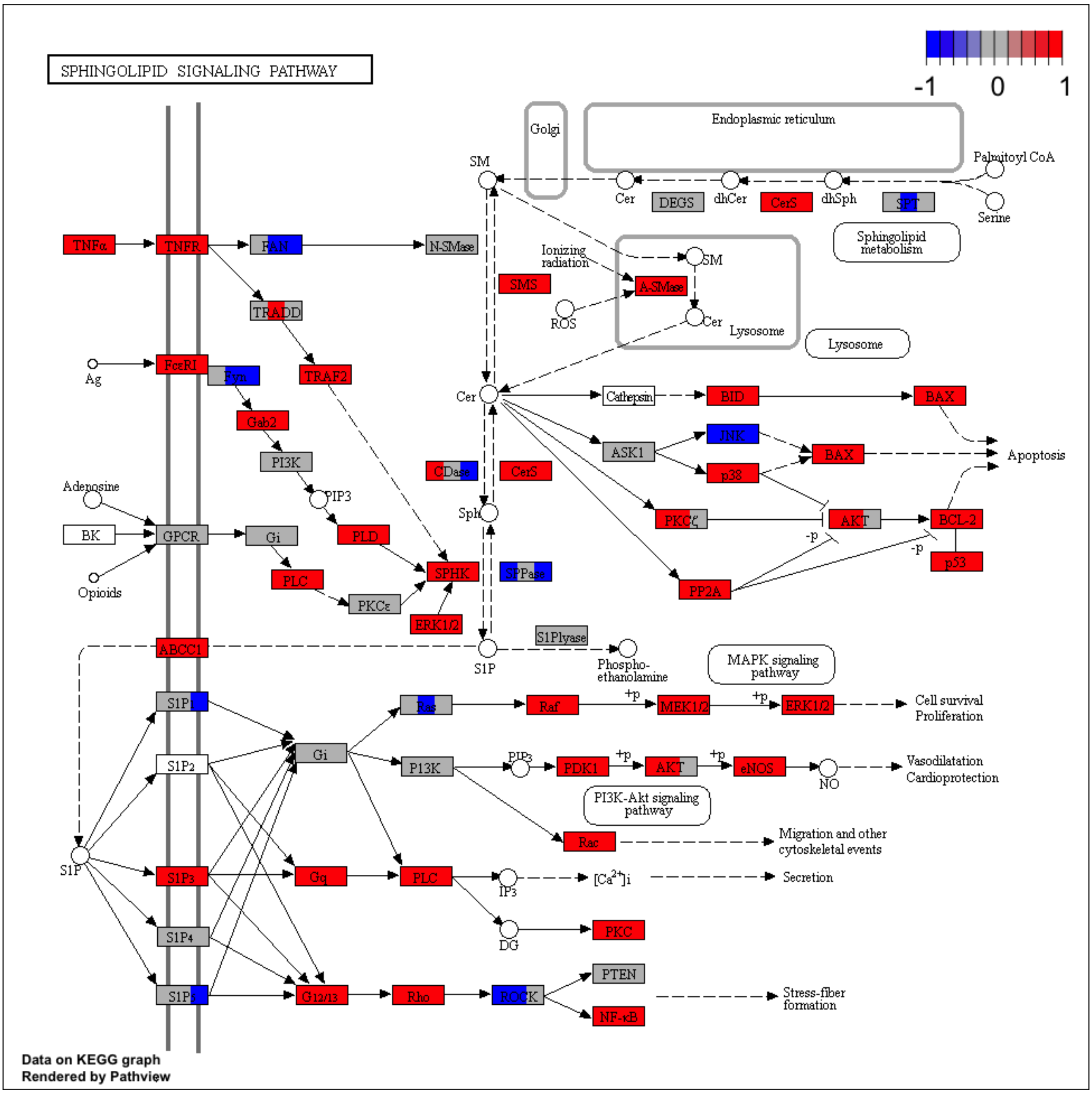

**Supplementary Figure S9: Sphingolipid signaling pathway.** Mapping of identified differentially expressed genes (DEGs) to sphingolipid pathways in the KEGG database. Rectangles show genes and the red and blue colors represent increased and decreased DEGs, respectively. Left, middle and right parts of the rectangles, respectively, show the results for subjects with fibrosis stage F0-1, F2, and F3-4, respectively.

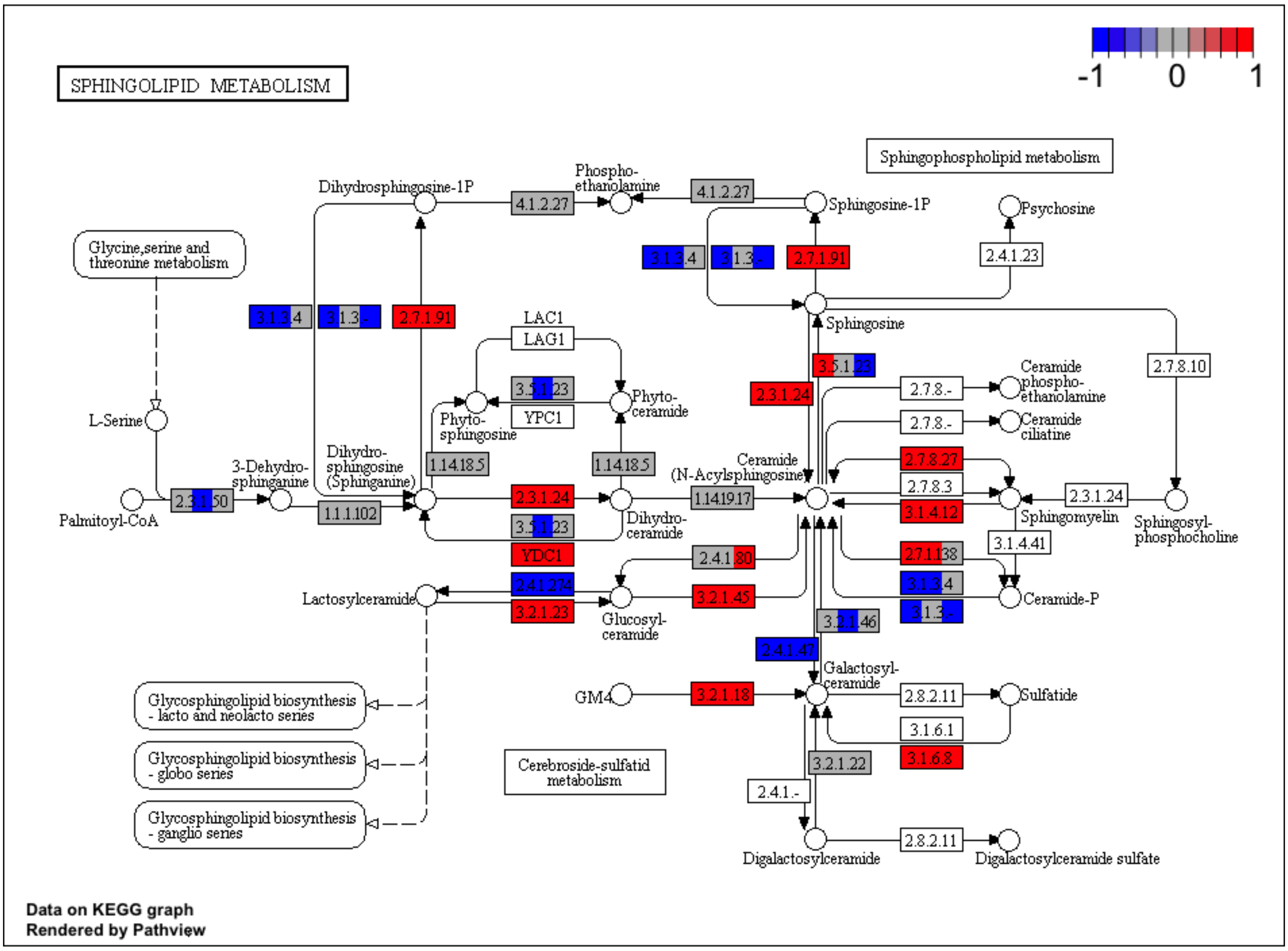

**Supplementary Figure S10: Sphingolipid metabolism pathway.** Mapping of identified differentially expressed genes (DEGs) to sphingolipid pathways in the KEGG database. Rectangles show genes and the red and blue colors represent increased and decreased DEGs, respectively. Left, middle and right parts of the rectangles, respectively, show the results for subjects with fibrosis stage F0-1 (K_[01]), F2 (K_2) or F3-4 (K_[34]), respectively.

#### Figures S11-S13: Expression of genes in specific synthesis and degradation pathways

Supplementary Figures S11-13 show the abundance of transcripts of genes in the sphingomyelin pathways. Boxplots shows gene abundance (counts per million; CPM) and heatmaps shows fold changes against mean value of the healthy control group (HP_0) for fibrosis stage F0-1 (K_[01]), F2 (K_2) and F3-4 (K_[34]) Results for low abundant genes excluded from the statistical analysis were shown as NA. Asterisks represent statistical significance against the HP_0 group, *: FDR < 0.05, **: FDR < 0.005, ***: FDR < 0.0005.

**a**

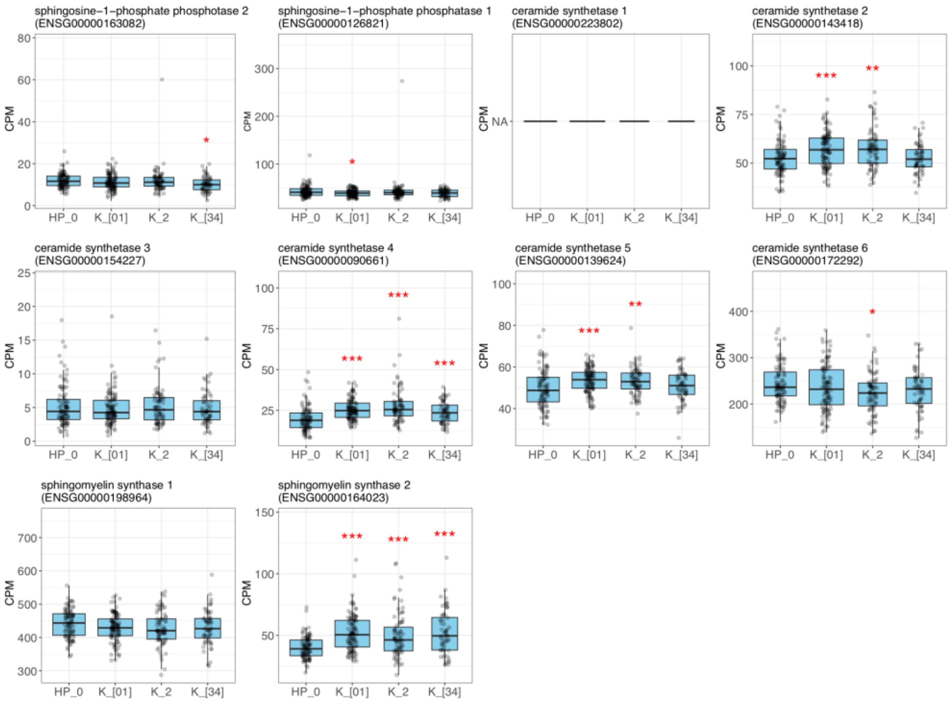

**b**

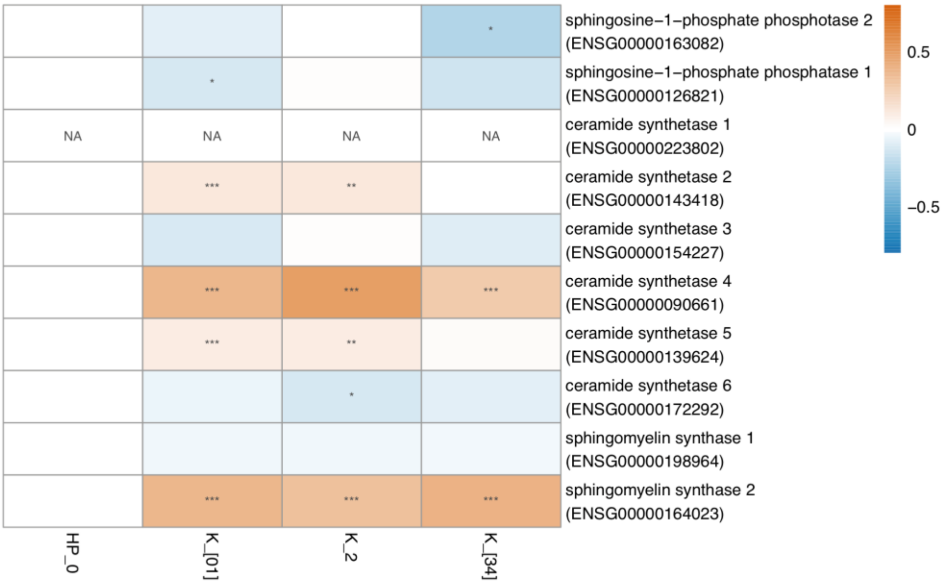

**Supplementary Figure S11: Biosynthesis pathway 1: sphingosine-1-p -> sphingomyelin.** Boxplots show gene abundance (counts per million; CPM), and heatmaps shows log2 fold changes against mean value of the healthy control (HP_0) group. Results for low abundant genes excluded from the statistical analysis were shown as NA. Asterisks represent statistical significance against the HP_0 group, *: FDR < 0.05, **: FDR < 0.005, ***: FDR < 0.0005. See text in the section for details. Fibrosis stage F0-1 (K_[01]), fibrosis stage F2 (K_2) and fibrosis stage F3-4 (K_[34]) using healthy matched controls as comparator (HP_0).

a

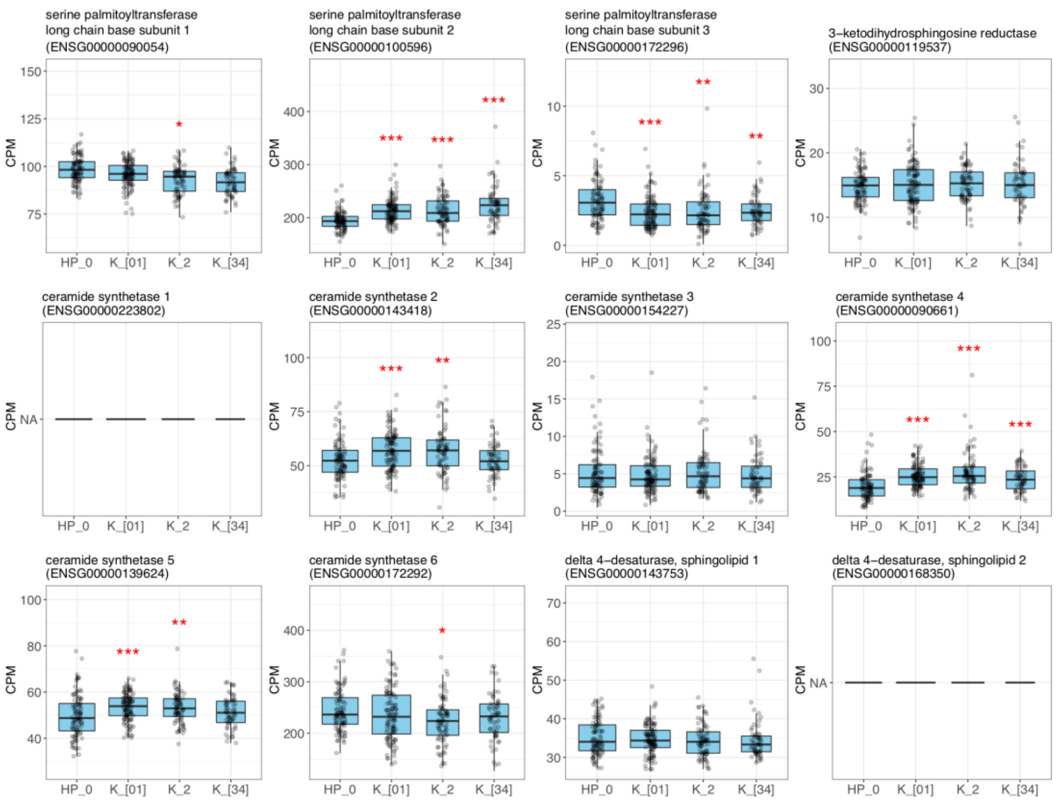

b

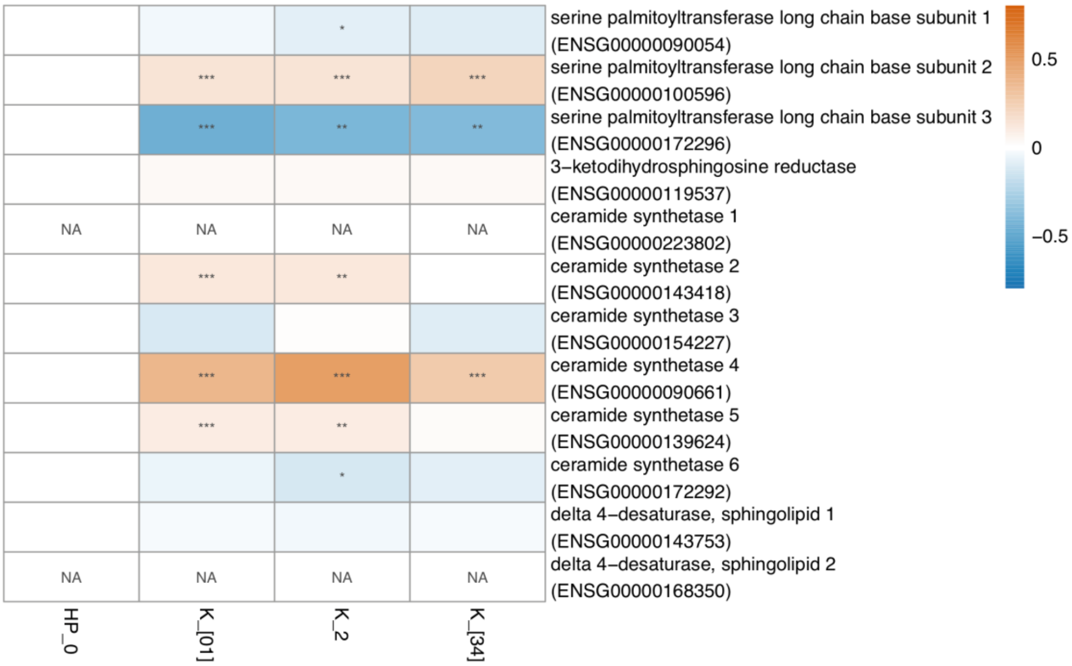

**Supplementary Figure S12: Biosynthesis pathway 2: serin + palmitoyl-CoA -> sphingomyelin.** See Supplementary Figure S11 for details.

**a**

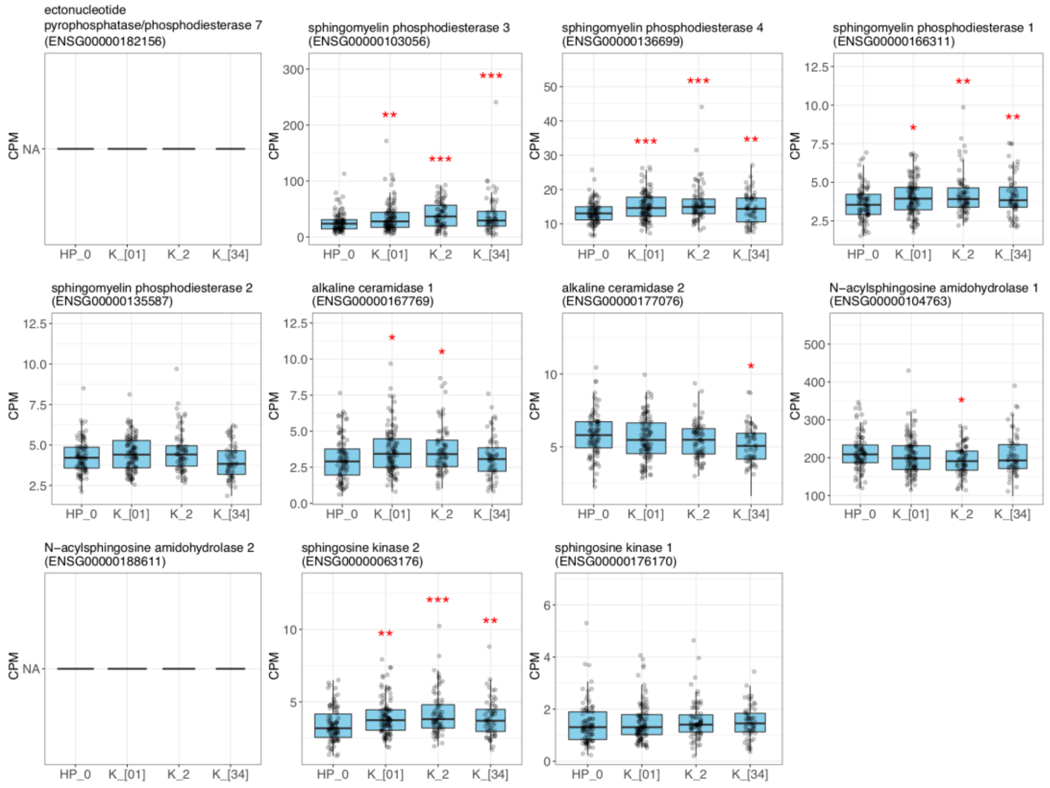

**b**

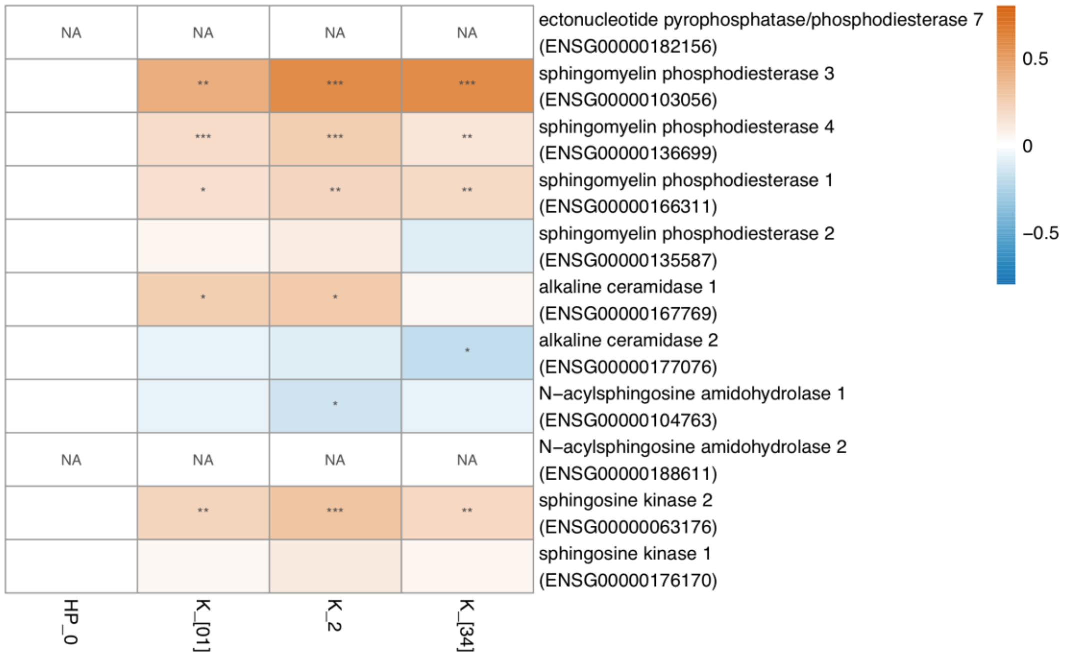

**Supplementary Figure S13: Degradation pathway: sphingomyelin -> sphingosine-1-p.** See Supplementary Figure S11 for details.

#### Causal Effects on Sphingomyelin Level

##### Effect of ALD on Blood Sphingomyelin Level

Effect: -0.03197
Standard error: 0.01158
p-value: 0.005788

The Mendelian randomization estimate to the causal effect is shown in Supplementary Figure S. Leave-one-out sensitivity analysis of the estimate is shown in Supplementary Figure S.

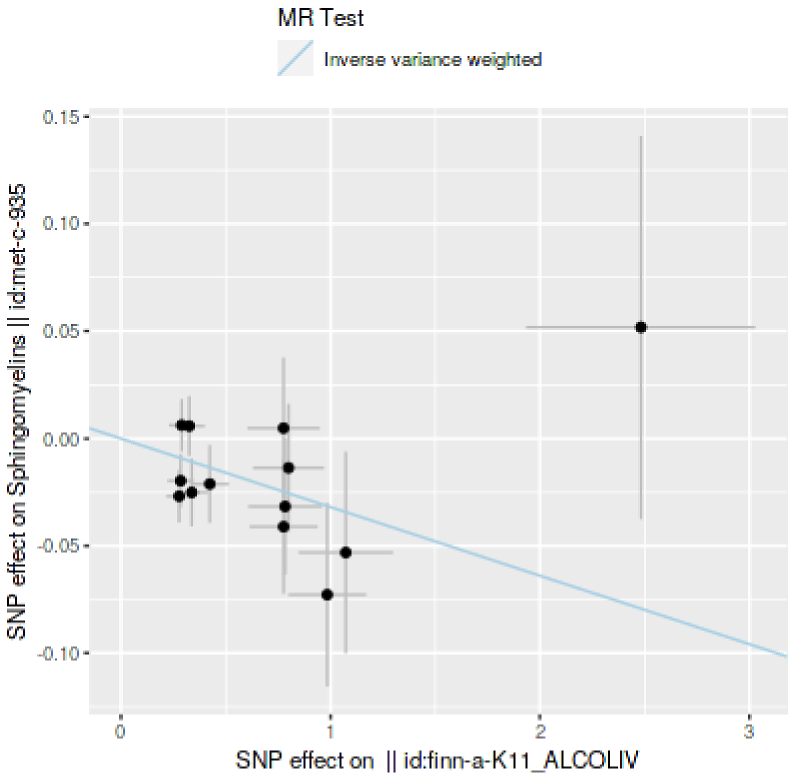

**Supplementary Figure S14: Causal effect of alcohol-related liver disease on blood sphingomyelin levels.** In two-sample Mendelian randomization meta-analysis, the single-nucleotide polymorphism (SNP) effect on blood sphingomyelin level (y-axis) is regressed against the SNP effect of alcohol-related liver disease (x-axis). Regression line (β = -0.032) that deviates from the horizontal level demonstrates the causal effect (p = 0.0058).

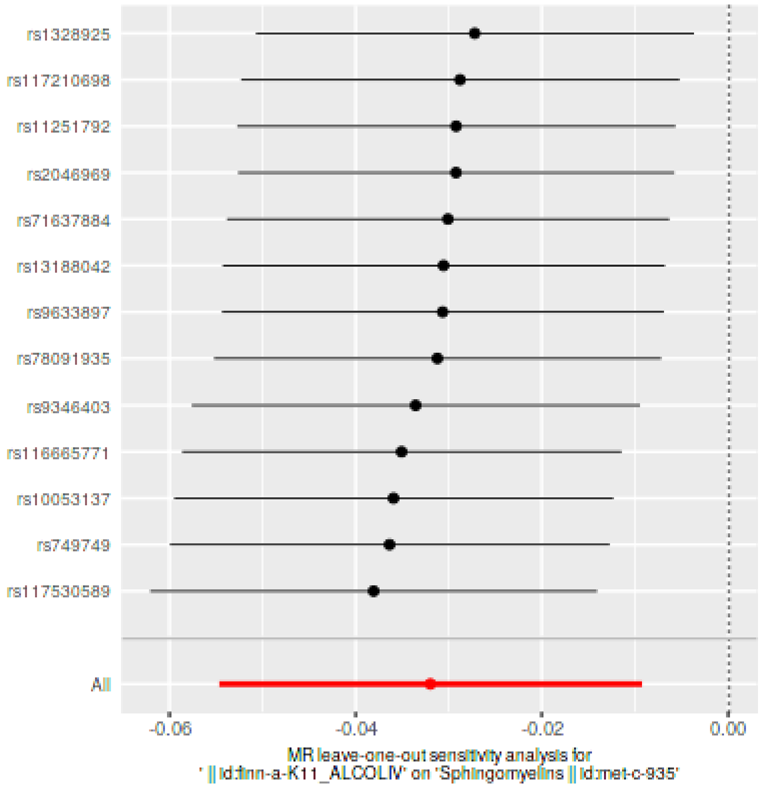

**Supplementary Figure S15: Leave-one-out sensitivity analysis to the Mendelian randomization (MR) analysis.** The MR-effect is shown, when each of the single-nucleotide polymorphisms (SNPs; y-axis), one at a time, are left out from the model.

##### Effect of NAFLD on Blood Sphingomyelin Level

Effect: 0.008137
Standard error: 0.0105
p-value: 0.4385

##### Effect of Alcohol Addition on Blood Sphingomyelin Level

Effect: 0.0089
Standard error: 0.05646
p-value: 0.8747

##### Effect of Blood Sphingomyelin Level on ALD

Effect: -0.1529
Standard error: 0.1854
p-value: 0.4094

#### Cellular Sphingolipid Metabolism

Figure S16: Cellular Sphingolipid Metabolism

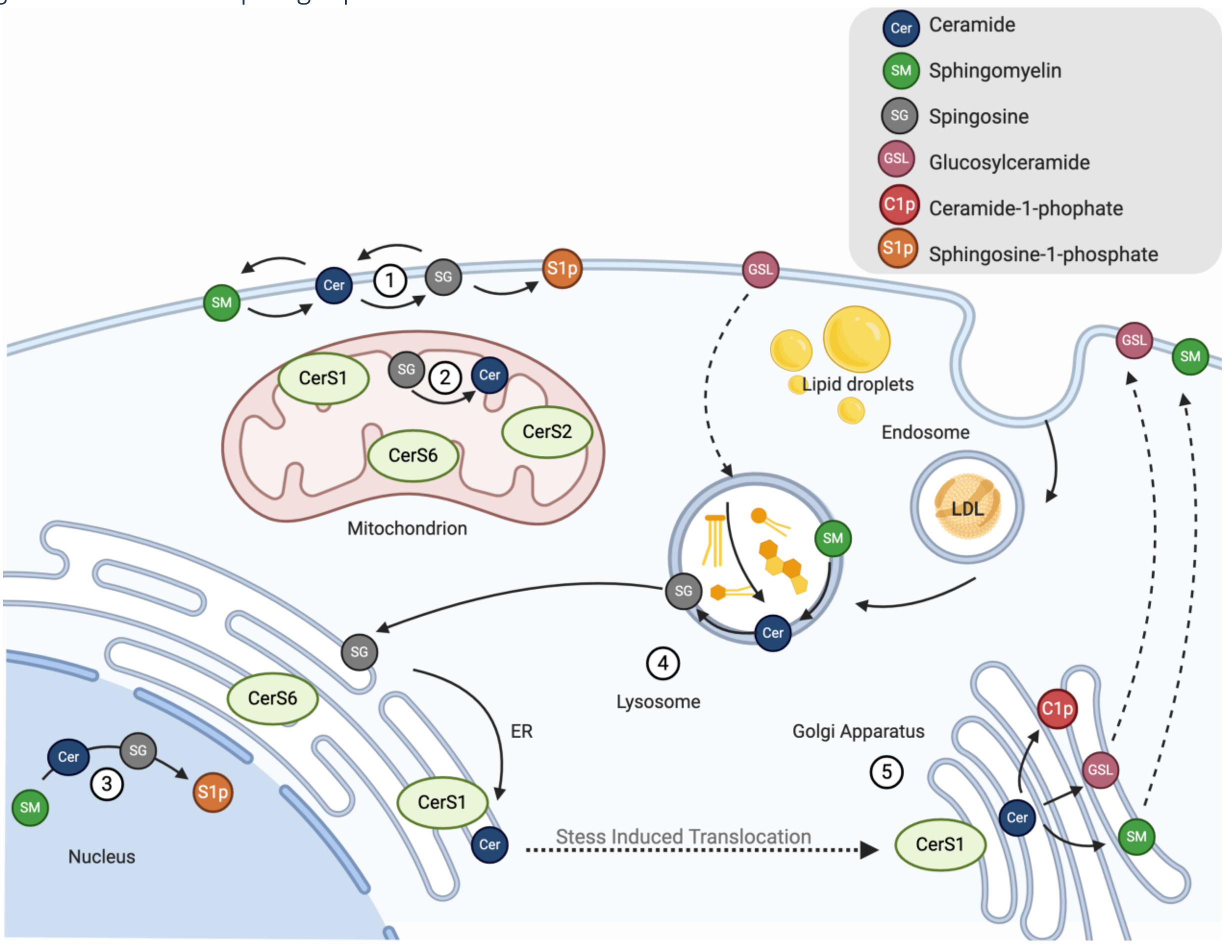

**Supplementary Figure S16: Cellular Sphingolipid Metabolism.** Possible Ceramide (Cer) and Sphingomyelin (SM) depletion pathways of known sphingolipid pathways in cells. Described as 1. Plasma membrane metabolism, 2. Mitochondria in which each ceramidase synthase (CerS1-6) catalyses different FA chain ceramides, 3. Nucleus metabolism leading to sphingosine-1-phospate, 4. Lysosomal digestion, 5. Ceramides can be translocated via vesicle-transport (dotted arrows) from the ER to the Golgi apparatus and further catalysed to ceramidase-1-phoshate. (Based on Turpin-Nolan et al. 2020.)
